# Duodenal microbiota and associated environmental enteric dysfunction in undernourished Bangladeshi women and gnotobiotic mice

**DOI:** 10.64898/2026.08.14.26360472

**Authors:** Kali M. Pruss, ZeNan L. Chang, Md. Shabab Hossain, M. Masudur Rahman, Mustafa Mahfuz, Reyan Coskun, Rumana Sharmin, A.H.M. Rezwan, Shafiqul Alam Sarker, Subhasish Das, Shah Mohammad Fahim, Md. Amran Gazi, Kelsey A. Hudson, Athziri Marcial Rodríguez, Haoxin Liu, Richard Kitchen, Alexandra E. Byrne, Clara Kao, Brooks B. Brodrick, Abbey Rose, Bishan Bhattarai, Darya Khantakova, Jose Fachi, Marco Colonna, Tahmeed Ahmed, Michael J. Barratt, Jeffrey I. Gordon

## Abstract

Undernutrition is an intergenerational global health challenge. Environmental enteric dysfunction (EED) is a small intestinal (SI) disorder characterized by villous atrophy, gut barrier dysfunction, malabsorption and systemic inflammation. To examine its pathogenesis and role in undernutrition, we performed esophagogastroduodenoscopy on undernourished Bangladeshi women with EED and their healthy counterparts. Histologic characterization of duodenal mucosal biopsies, aptamer-based proteomic analyses of their duodenal mucosa and plasma, plus metagenomic analyses of their duodenal and fecal microbiota, revealed associations between bacterial taxa and duodenal tissue and plasma proteomes indicative of EED. Colonization of germ-free female mice with consortia of duodenal bacteria from these women, followed by measurements of SI bacterial abundances, SI cellular patterns of gene expression (single nucleus RNA-seq), plus proteomic and flow cytometric analyses disclosed bacterial, epithelial, and immune features in dams and their offspring resembling those in the women. These findings have diagnostic and therapeutic implications.

## INTRODUCTION

Undernutrition in children, defined anthropometrically by impaired linear growth (stunting) and/or by impaired ponderal growth (wasting) affects ∼200 million children under age five globally, with the greatest burden of disease in the global South^1–4^. Moreover, undernutrition is an intergenerational problem with maternal undernutrition being a key predictor of undernutrition in their offspring^4–6^. Environmental enteric dysfunction (EED) is a subclinical small intestinal disorder characterized by villous blunting with accompanying reduction in epithelial nutrient absorptive area, mucosal barrier dysfunction and local as well as systemic inflammation^7–13^. The true prevalence of EED, its pathogenesis and its contribution to intergenerational undernutrition remain poorly understood, in part due to a paucity of serum and fecal biomarkers validated by concurrent histopathologic examination of small intestinal mucosal biopsies^7^.

We previously studied a cohort of children residing in an urban slum located in the Mirpur district of Dhaka, Bangladesh who were stunted or at risk for stunting and failed a standard nutritional intervention for linear growth faltering^14,15^. These children were part of the Bangladeshi EED study (BEED) [ClinicalTrials.gov number, NCT02812615], where informed consent was obtained for esophagogastroduodenoscopy (EGD) to determine whether their duodenal mucosal biopsies showed evidence of EED and where their duodenal microbiota and duodenal mucosal proteome could be characterized. 94% (85/90) of these children had histopathologic evidence of EED^14^. For ethical reasons, EGD could not be performed on healthy children living in the same locale.

We cultured bacteria from the duodenal aspirates obtained from children in the BEED study and assembled distinct consortia that were introduced into germ-free adult female^16^. Subsequent dam-to-pup transmission of these consortia results revealed how components of the duodenal microbiota associated with EED can produce local and systemic inflammatory responses in adult female gnotobiotic mice as well as impaired prenatal and postnatal growth in their offspring. Moreover, signaling pathways related to intestinal epithelial cell renewal, barrier integrity and immune function were altered in pups^16^. This provided preclinical evidence for a causal relationship between the duodenal microbiota and EED in children.

EED in adults has been understudied, particularly in women of reproductive age. A recent case-control observational study described the histopathology of duodenal mucosal biopsies obtained at the time of EGD of undernourished (BMI<18.5 kg/m^2^) adult women residing in Mirpur; 75% exhibited histopathological evidence of EED, including villus atrophy, crypt hyperplasia and immunoinflammatory changes^17–19^. The availability of a cohort of healthy women living in the same locale as those who were undernourished provided an opportunity to address questions related to the pathogenesis of EED. Specifically, are there duodenal and plasma proteomic features that are discriminatory for undernourished women with histopathologic evidence of EED versus their healthy counterparts? How do these features correlate with the representation of microbial taxa in their duodenal and fecal microbiota? Can gnotobiotic mice be used to test whether observed microbial features are causally related to EED? Finally, to what extent are these discriminatory features also found in children with EED?

To address these questions, in the current study we perform EGD on cohorts of well-nourished Bangladeshi women living in Dhaka, determined the extent to which their duodenal biopsies had histopathologic features of EED, and then compared their duodenal mucosal and plasma proteomes as well as their duodenal and fecal microbiota to those of undernourished women with EED. Application of multi-omic analytic methods to these biospecimens allowed us to identify significant associations between the abundances of duodenal bacterial taxa and duodenal proteins that distinguished women with EED from their healthy counterparts as well as candidate disease biomarkers present in their plasma. Bacteria cultured from duodenal aspirates obtained from these undernourished and well-nourished women were used to colonize germ-free adult female mice. Transcriptional and proteomic analyses of adult female mice and their offspring revealed links between duodenal bacteria and enteropathy as well as growth faltering that were reflective of EED in women and children. Finally, duodenal bacteria that were associated with immuno-inflammatory responses in our gnotobiotic mouse experiments and in the duodenal mucosal proteome of women were compared to analogous samples from children with EED to further refine our understanding of organisms associated with enteropathy.

## RESULTS

### Study design

The design, recruitment, and exclusion criteria for women living in Mirpur for the BEED study are described in detail elsewhere^15,17^. Briefly, undernourished (BMI<18.5 kg/m^2^) women, 18-45 years old, were recruited for nutritional therapy. Those whose BMI failed to improve (<10% increase) after three months of the nutritional intervention (see *Methods*) underwent EGD for collection of duodenal mucosal biopsies and duodenal luminal aspirates. Fecal samples and plasma were collected concurrently from 22 of these women (**Supplementary Table 1a**). As part of the BEED study, comparably aged, well-nourished women (BMI=18.5-30 kg/m^2^) residing in Mirpur who were seeking treatment for dyspepsia and were eligible for EGD underwent the procedure, had normal endoscopic findings, and thus were diagnosed as having functional dyspepsia. Duodenal biopsies and aspirates were collected from these individuals during EGD (n=33, **Fig. 1a,b**). Only one of these 33 women had no histopathologic evidence of EED (defined as a score of 0 according to the grading scale described in *Methods* and refs.^17,18^, **Supplementary Table 1a**). Therefore, we recruited a second cohort of well-nourished women from Dhaka who were also 18–45 years old. They had BMIs of 20-25 kg/m^2^ and had sought treatment for functional dyspepsia. These women (n=25) underwent EGD and were diagnosed with functional dyspepsia as above. Duodenal aspirates and mucosal biopsies were collected during EGD with concurrent collection of plasma and fecal specimens; 19 of these indivdiuals had biopsies with histopathology score of 0 (**Supplementary Table 1a**). Members of the low BMI cohort with histopathologic evidence of EED, and the two well-nourished cohorts (with or without evidence of EED) were characterized using plasma proteomics (SomaScan 11k arrays), duodenal mucosal proteomics (SomaScan 7k arrays), and duodenal and fecal microbiota profiling (full-length bacterial 16S rRNA amplicon sequencing).

**Fig. 1.**
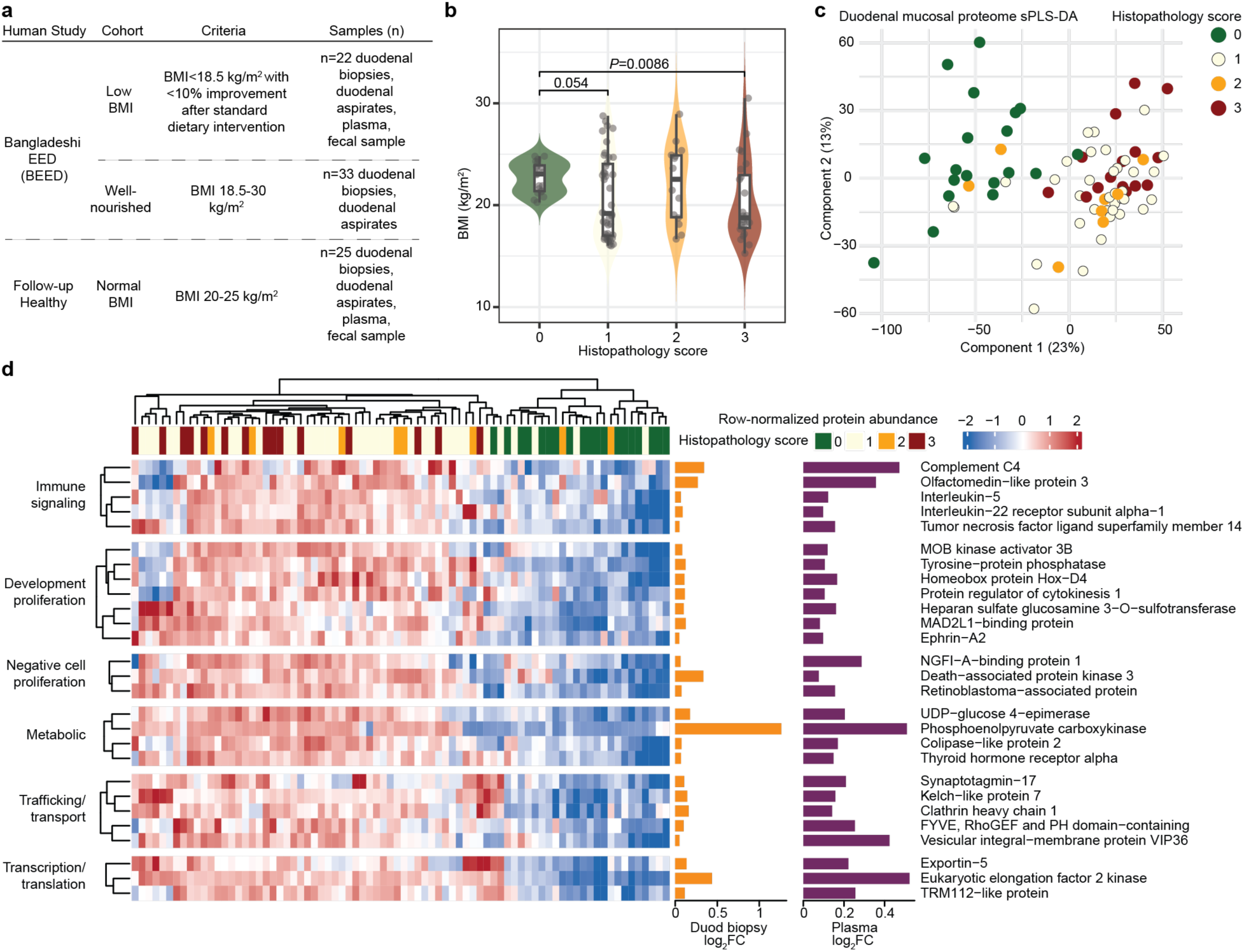
Undernutrition in adult Bangladeshi women with histopathologic evidence of EED is characterized by intestinal and systemic inflammation. **a,** Human studies investigating the microbial contribution to undernutrition in adult women; overview of human biospecimens collected. **b,** Distribution of BMI for participants within each histopathology score (Kruskal Wallis *P*=0.067, *P*-values shown are the result of Wilcoxon rank-sum tests). **c,** Dimensionality reduction of of duodenal mucosal proteomes (n=78 women, sparse partial least-squares discriminant analysis). Color denotes histopathology score. **d,** A subset of the 34 proteins whose levels were significantly higher in women with EED compared to healthy controls in duodenal mucosal biopsies and plasma (*limma* adjusted *P*-value<0.1). The heatmap depicts log_10_-transformed, row-normalized levels in the duodenal mucosa. Bar plots show the magnitude of differences between women with and without EED in biopsies and plasma. See Supplementary Table 1d for a full list of protein annotations and their thematic groupings.

### Duodenal and plasma proteomes of women with EED

#### Duodenal proteome

Dimensionality reduction (sparse partial least squares discriminant analysis, sPLS-DA)^20^ showed that protein composition within the duodenal mucosa separated primarily by histopathology score (**Fig. 1c**). Across all three cohorts, levels of 1,375 duodenal proteins were significantly different between women with EED (860 proteins higher in n=59 women with histopathology score 1-3) compared to those without EED (521 proteins higher in biopsies of n=19 women with histopathology score 0, *limma* adjusted *P*-value<0.1; **Supplementary Table 1b(i)**). Twenty-two Reactome pathways were significantly enriched in the duodenal mucosal proteomes of women with EED; they included several pathways related to bacterial recognition and response (‘Toll Like Receptor Cascades’, ‘MyD88 cascade’, ‘TRAF6 mediated induction of NFkB and MAPK’ **Extended Data Fig. 1a, Supplementary Table 1b(ii)**).

#### Plasma proteome

A total of 517 plasma proteins was significantly elevated in undernourished women with EED (n=22 women with histopathology score ≥ 1); 191 proteins were elevated in healthy controls without EED (n=19 women with histopathology score 0; *limma* P-adj < 0.1, **Supplementary Table 1c, Extended Data Fig. 1b**). Plasma proteins elevated in EED included complement components (C3, C4A/B), pro-inflammatory cytokines and their receptors (IL-5, IL-21, IL-36A, IL22RA1, IFNA6, IFNA10, Macrophage migration inhibitory factor MIF) and the bacterial sensing protein TLR1 (**Supplementary Table 1c**).

To identify potential biomarkers of EED, we cross-referenced the set of duodenal mucosal proteins elevated in women with EED (histopathology score ≥1, 860 proteins) with plasma proteins elevated in undernourished women with EED (517 proteins): 34 proteins satisfied these criteria. **Fig. 1d** and **Supplementary Table 1d** describe their functions, which include immune signaling, bacterial sensing, cell adhesion, regulation of cell proliferation and mitochondrial function.

### Associations between duodenal bacterial strain abundances and duodenal mucosal proteins

We sequenced full-length 16S rRNA amplicons generated from DNA isolated from duodenal aspirates recovered from women with (n=60) and without (n=20) histopathologic evidence of EED (**Extended Data Fig. 1d**). The absolute abundances of 38 bacterial taxa significantly increased or decreased with histopathology score (zero-inflated Gaussian mixture model *P*-adj<0.1; *Methods*); they included organisms previously associated with the oral microbiota (**Fig. 2a, Supplementary Table 2a-c**; see ***Supplementary Results*** and **Supplementary Table 3a,b** for a supervised learning analysis identifying bacteria in duodenal aspirates and proteins in duodenal tissue biopsies that together predicted disease status).

**Fig. 2.**
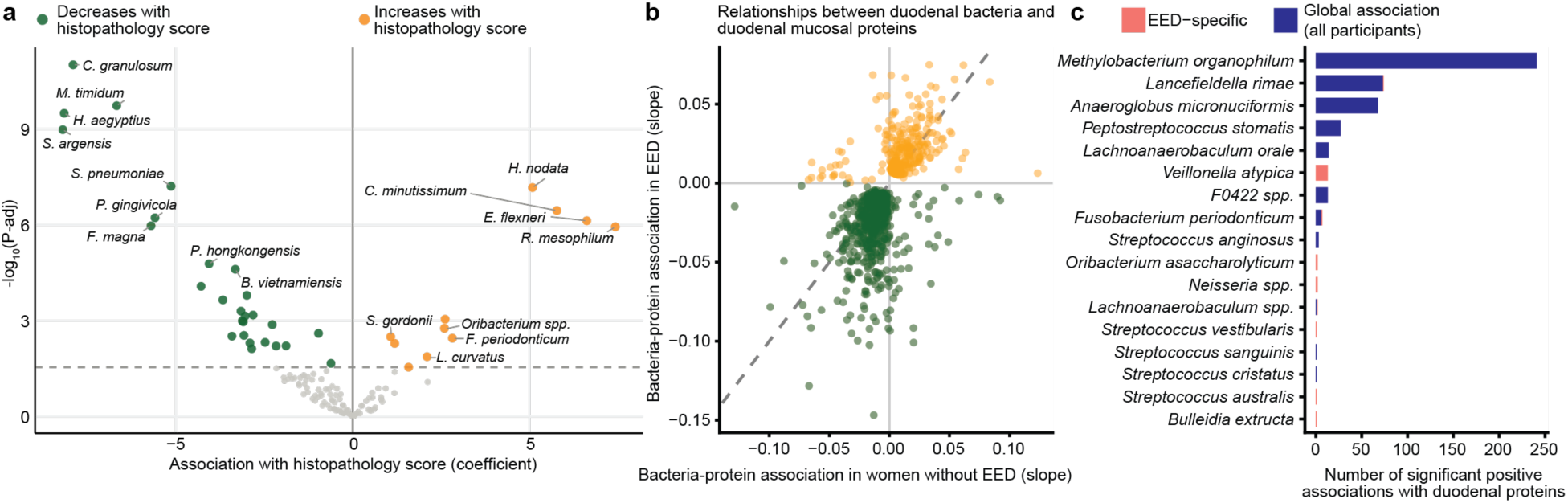
Duodenal microbiota and its associations with the duodenal mucosal proteome in women with EED. **a,** Bacteria whose absolute abundances in duodenal aspirates increased (positive, orange) or decreased (negative, green) with histopathology score (zero-inflated Gaussian mixture model). See **Supplementary Table 2b** for full list of bacteria including genus and species. **b,** ß coefficients for linear models relating bacterial abundance in duodenal aspirates to duodenal mucosal protein levels. Bacteria-protein linear relationships that were significant are included and are colored according to the direction of their association in women with EED (orange: positive, green, negative). **c, D**uodenal bacterial taxa whose abundances exhibited the greatest number of positive associations with duodenal mucosal proteins in all participants (global association) or in women with EED, but not in women without EED (‘EED-specific’, see also **Extended Data Fig. 2a**). For panel **a**, n=80 women, for panels (**b,c**), n=78 women.

To identify duodenal bacteria associated with the duodenal mucosal proteins, we next performed linear regressions on the absolute abundances of bacterial taxa in duodenal aspirates with levels of proteins in matched biopsies (**Fig. 2b**, **Extended Data Fig. 2a**). The bacterial taxa whose abundances exhibited the greatest number of significant positive associations with duodenal mucosal proteins were *Methylobacterium organophilum* followed by *Lancefieldella rimae* and *Anaeroglobus micronuciformis* (**Fig. 2c**). The absolute abundances of these and other duodenal bacteria were positively linearly related with levels of a large set of innate immune signaling proteins (**Extended Data Fig. 2b**), including Sialic Acid-binding Ig-like Lectins (SIGLEC14), Interleukins (IL22, IL25) and their receptors (IL11RA, IL17RC, IL22RA2), Interferons (Interferon (IFN)A7, IFNL2, IFNLR1), components of the complement system (C1Q, C1QL3, C3, C5, C6), pathogen sensors and defensins (Regenerating Family Member (REG) 1B, REG3B; Defensing (DEF)6, DEFA5, DEFB110, DEFB115, DEFB116, DEFB129), proteins in cell death pathways (Caspases 14 and 10, NLR family pyrin domain containing 1 (NLRP1)) and chemokines (C-C Motif Chemokine Ligand (CCL)7, CCL2; C-X-C Motif Chemokine Ligand 2, CXCL2, Granzyme B, GZMB) (**Supplementary Table 3c**). A functional enrichment analysis disclosed that these proteins are associated with leukocyte migration, cytokine signaling, cell death and reactive oxygen species detoxification (**Extended Data Fig. 2b, Supplementary Table 3d**). Conversely, functions enriched amongst duodenal mucosal proteins whose levels were significantly negatively associated with the abundances of duodenal bacterial taxa included several metabolic pathways (‘Cholesterol transport and metabolism’, ‘Lipid metabolism’, ‘Amino acid metabolism’) as well as components of pathways involved in responses to steroid hormones (see **Supplementary Table 3e** for details).

A subset of these bacteria-protein pairs exhibited significant differences between women with and without histopathologic evidence of EED (**Extended Data Fig. 2a,c, Supplementary Table 3c**). Duodenal mucosal proteins that were significantly positively associated with bacterial taxa in women with EED but not in women without EED were involved in inflammation, barrier defense (Granzyme B, GZMB; Leucine rich repeat-containing 15, LRRC15; General transcription factor 2i, GTF2I; Prolactin-inducible protein. PIP; Serum amyloid A1, SAA1) and detoxification of lipid aldehydes/reactive carbonyls (Aldehyde dehydrogenase 7 family member A1, ALDH7A1; Carbonyl reductase 3, CBR3).

#### Fecal microbiota

Long read sequencing of full-length bacterial 16S rRNA amplicons identified 538 unique species in fecal samples from members of the Low BMI (n=38) and Normal BMI (n=25) cohorts (**Supplementary Table 4a**). After prevalence filtering (taxa present in ≥ 9 women), 200 unique species remained and were included in subsequent analysis. The relative abundances of 83 taxa were significantly associated with histopathology score (**Extended Data Fig. 4a**, **Supplementary Table 4b**, adjusted *P-*value<0.05). A sparse partial least squares-discriminant analysis (sPLS-DA) identified fecal taxa that were predictive of EED (**Extended Data Fig. 4b,c**). In addition, we quantified common enteropathogens in DNA extracted from fecal samples using a microfluidics-based quantitative PCR platform^21^; *Giardia lamblia* was significantly more prevalent in women with EED (**Extended Data Fig. 4d, Supplementary Table 4c**), consistent with previous reports in other populations suggesting that subclinical pathogen exposure contributes to pathophysiology of EED and undernutrition^22–25^.

### Identifying inflammation-inducing duodenal bacteria from adult women

We used gnotobiotic mice to identify members of the duodenal microbiota from women with EED that could produce mucosal and systemic inflammation. To do so, we cultured bacteria from duodenal aspirates under anaerobic and microaerophilic conditions. A total of 480 isolates representing 53 unique species were obtained from 14 undernourished women with EED and 125 isolates representing 35 unique species from 11 well-nourished women (BMI 19-25 kg/m^2^) (species identified from sequencing full length 16S rRNA gene amplicons, **Extended Data Fig. 3**, **Supplementary Table 5a,b**). Isolates from undernourished or healthy women were pooled and gavaged to just-weaned mice fed a diet representative of that consumed by women living in Mirpur, Bangladesh (‘Adult Mirpur’) where the BEED clinical study had been performed. To focus on colonizing members that demonstrated a capacity to elicit disparate levels of inflammation, cecal contents from animals harboring (i) the EED-donors’ cultured bacterial consortium that had the highest levels of serum biomarkers of EED (complement C3 as identified above, and LCN2 and MMP8; previously identified in children with EED and also found to be elevated in gnotobiotic mouse models of the children’s duodenal microbiota^14,16^) and (ii) the well-nourished donors’ consortium with the lowest levels of these three host proteins, were each transferred to separate groups of naïve germ-free recipients (**Supplementary Table 6a**). After an 18-day colonization, mice harboring the ‘inflammatory’ consortium from undernourished women exhibited significantly elevated levels of serum LCN2 (**Extended Data Fig. 5a**), C3 (**Extended Data Fig. 5b**), IL-17A (**Extended Data Fig. 5c**) and IL-6 (**Extended Data Fig. 5d**), as well as reduced weight gain (**Extended Data Fig. 5e**, **Supplementary Table 6a,b**) compared to recipients of the well-nourished donors’ consortium. We refer to these two consortia as the ‘aSI-L’ (adult small intestinal-Low BMI) and ‘aSI-H’ (Healthy BMI) consortia (see *Methods*).

Duodenal bacteria from women with EED that were associated with inflammation in these mice (i.e. significantly higher in aSI-L compared to aSI-H animals) included *Ligilactobacillus salivarius, Streptococcus gordonii, Streptococcus infantarius, Granulicatella adiacens, Haemophilus parainfluenzae, Escherichia flexneri* and *Klebsiella granulomatis* (**Extended Data Fig. 5f**, **Supplementary Table 6c,d**).

#### Transcriptional responses of small intestinal epithelial cell lineages to EED versus healthy bacterial consortia

Duodenal, jejunal, and ileal intestinal segments were collected from adult female mice fed the ‘Adult Mirpur’ diet that had been colonized for >5 weeks with the aSI-L or aSI-H consortia (see *‘Tissue collection from adult female mice’* in *Methods* for how these segments were operationally defined). No statistically significant differences in villus length or crypt depth were observed along the length of the small intestines between animals belonging to the two treatment groups (**Supplementary Table 7a**).

We performed single nucleus RNA-Sequencing (snRNA-Seq) to assess intestinal cellular responses to the aSI-L compared to aSI-H consortium. A total of 45,048, 47,461 and 36,441 nuclei were collected from three intestinal segments (duodenum, jejunum, and ileum; n=4 mice/group, see *Methods* for quality control). The resulting transcriptomes were categorized into epithelial and mesenchymal cell clusters (lineages) based on marker gene expression (**Fig. 3a**). Enterocytes were organized into subclusters representing subpopulations positioned at different locations along the crypt-villus axis^26,27^.

**Fig. 3.**
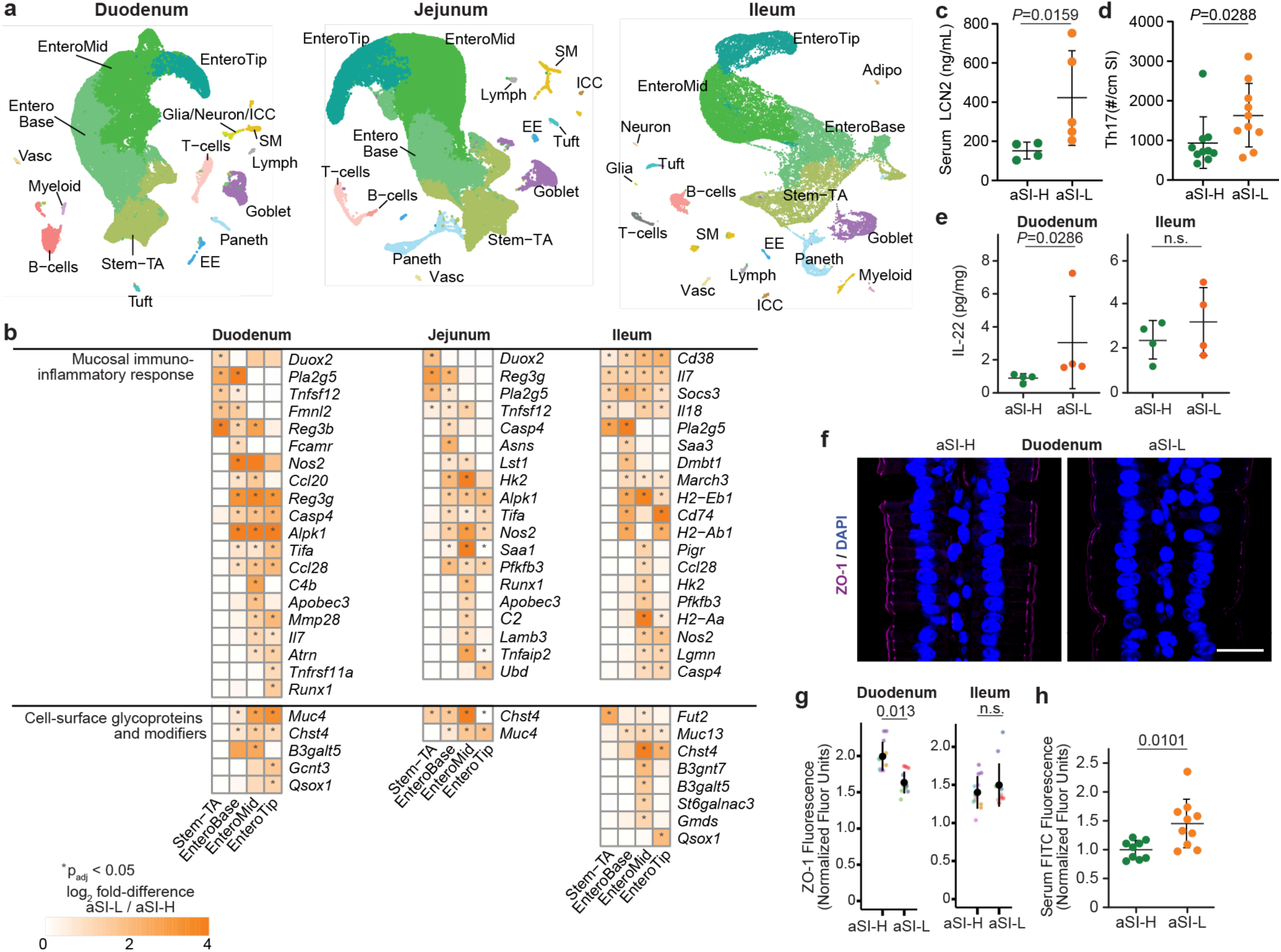
Duodenal bacteria cultured from Low BMI women with EED impair gut barrier function and induce inflammatory epithelial signaling in adult female mice. **a,** Uniform Manifold Approximation and Projection (UMAP) dimensionality reduction of snRNA-Sequencing in duodenal, jejunal, and ileal tissue from aSI-L and aSI-H adult female mice (n=4 mice/intestinal segment/group). Abbreviations: Stem-TA, intestinal stem cell and transit amplifying cells; EnteroBase, EnteroMid, EnteroTip, enterocytes from villus base, mid-villus, and villus tip; EE, enteroendocrine; ICC, interstitial cells of Cajal; Lymph, lymphatic endothelial cells; SM, smooth muscle; Vasc, vascular endothelial cells; Adipo, adipocytes. **b,** Genes involved in mucosal immuno-inflammatory responses as well as genes encoding cell surface glycoproteins and their modifiers whose expression was significantly higher in intestinal epithelial nuclei of aSI-L compared to aSI-H adult female mice (*DESeq2* Wald test with Benjamini-Hochberg correction, ‘pseudo-bulk’ differential expression). **c,** Serum LCN2 levels (n=4-5 mice/group, bars denote means). **d,** Proportions of T cells in the small intestinal lamina propria (n=10 mice/group, bars denote means). **e,** Levels of IL-22 in homogenized duodenal and ileal tissue (n=4 mice/group, bars denote means). **f,** Representative immunofluorescent images for staining of zonula occludens-1 (ZO-1) in the duodenum of adult female mice colonized with the aSI-L or aSI-H consortia (scale bars=8 μm). **g,** Mean fluorescence intensity (n=3 villi/segment/mouse for n=4 mice/group, mean ± s.d. shown). A linear mixed-effects model *(Measure ∼ Microbiota + (1 | Mouse ID)*) was used to assess statistical significance; each color denotes a different mouse. h, FITC-4 kDa dextran in serum 4 hours after gastric gavage (n=9-10 mice/group, bars denote means). For **c,d,e,** and **h,** Wilcoxon rank-sum tests.

‘Pseudo-bulk’ analysis of epithelial cell clusters revealed statistically significant elevations in transcripts related to inflammation in aSI-L adult females compared to aSI-H (**Fig. 3b**; see **Supplementary Table 7b-d** for full lists of differentially expressed genes across cell clusters in each small intestinal segment and along the crypt-to-villus axis). Genes more highly expressed in enterocytes of aSI-L animals included (i) those with antimicrobial activities (inducible nitric oxide synthase, *Nos2*; dual oxidase 2, *Duox2*; a secreted calcium-dependent phospholipase A2, *Pla2g5*; regenerating islet-derived protein 3 *Reg3b* and *Reg3g*), (ii) microbial pattern recognition receptors (alpha-protein kinase 1, *Alpk1* and its signaling adaptor TNF receptor-associated factor-interacting protein with forkhead-associated domain, *Tifa*), (iii) genes involved in responses to inflammation (an inflammatory caspase, *Casp4*; the antiviral innate immune effector apolipoprotein B mRNA-editing enzyme catalytic polypeptide-like 3, *Apobec3*; glycolytic genes *Hk2* and *Pfkfb3*; serum amyloid A, *Saa*), (iv) chemokines involved in homing of lymphocytes to the mucosa (*Ccl28* and *Ccl20*); (v) MHCII complex components (*H2-Aa*, *H2-Ab1*, *H2-Eb1*, *Cd74*) and a peptide processing enzyme for MHCII presentation (*Lgmn*); and (vi) genes related to the mucus barrier (cell surface mucins *Muc4* and *Muc13*; mucin-modifying enzymes *Qsox1, B3galt5*, *B3gnt7*, *Chst4*, *Fut2*, *Gcnt3*, *St6galnac3*, **Fig. 3b**).

Cross-referencing the protein products of genes whose expression was significantly elevated in the duodenum of aSI-L mice with duodenal proteins that were significantly higher in women with EED revealed shared elevations involving metabolic changes associated with inflammation (*Hk2*, *Pfkfb4*), myeloid cell survival and proliferation (*Il34*), and regulation of IL-22/STAT3 signaling (*Ptk6*) (**Extended Data Fig. 6a,b**).

### Immune cell profiling and assay of mucosal barrier function

A number of genes whose expression was significantly elevated in enterocytes in aSI-L adult females were previously shown to be upregulated in intestinal epithelial cells in response to Th17-cell induction in conventionally-raised mice (*e.g. Duox2, Fut2, Reg3b*, *Reg3g, Saa1*, *Saa3*)^28^ and/or upregulated upon exposure of an intestinal cell line to IL-22 which is produced by Th17 cells (*e.g. B3gnt7*, *Dmbt1*, *Duox2*, *Fut2*, *Gcnt3*, *Hk2*, *Nos2*, *Socs3*, *Tifa*, *Ubd*)^29^. To assess immune cell representation in the small intestinal lamina propria, we colonized additional groups of germ-free adult female mice fed the Adult Mirpur diet by horizontal transmission (coprophagia) from previously colonized aSI-L and aSI-H mice (the same groups used for snRNA-Seq analyses). The resulting aSI-L mice had significantly elevated serum levels of LCN2 compared to their aSI-H counterparts (**Fig. 3c, Supplementary Table 7e**). Flow cytometry of lymphocytic and myeloid populations demonstrated significant elevations in the absolute numbers of Th17 cells in the small intestinal lamina propria of aSI-L mice and elevations in the percentages of Th17, γδT, and RORγt^+^ regulatory T cells (**Fig. 3d, Supplementary Table 7f,g**) – consistent with the Th17-associated enterocyte transcriptional responses described above.

Duodenal goblet cells in aSI-L female mice exhibited significant reductions in expression of *Zg16* (**Supplementary Table 7b**), a secreted protein with lectin domains that can aggregate Gram-positive bacteria to limit their penetrance through the mucus layer^30^. Levels of IL-22 protein, which was previously shown to reduce ZO-1 expression in intestinal cells^31^, were significantly higher in homogenized duodenal tissue of aSI-L adult females (**Fig. 3e**). Consistent with these findings, immunostaining of proximal SI enterocytes for the tight junction protein zonula occludens-1 (ZO-1) demonstrated a reduction in aSI-L compared to aSI-H animals (**Fig. 3f,g, Supplementary Table 7e**). Based on these observations, we directly measured gut permeability by gastric gavage of FITC-4kDa dextran in aSI-L and aSI-H mice. A significant increase in passage of FITC-dextran into the serum of aSI-L mice indicated elevated gut permeability (**Fig. 3h, Supplementary Table 7h**). Together, these data suggest that the aSI-L consortium imparts a Th17 intestinal mucosal immune response associated with impaired gut barrier integrity in adult female gnotobiotic mice.

### Intergenerational (dam-to-pup) transmission of the aSI-L and aSI-H consortia

To explore potential consequences of intergenerational transmission of the aSI-L and aSI-H bacterial consortia on offspring, we colonized a separate group of adult female and male germ-free mice fed the Adult Mirpur diet with either the aSI-L or aSI-H consortium for one week prior to onset of mating. Their offspring were weaned onto the Mirpur-18 diet and euthanized on postnatal day 37 (P37) (n=35 offspring/group; **Fig. 4a**). Post-weaning weight gain was significantly reduced in P37 offspring of dams harboring the aSI-L consortium (**Fig. 4b**). Microcomputed tomography of their femurs revealed that the reduced weight gain in aSI-L mice was accompanied by reduced trabecular bone mineral density and increased cortical bone porosity compared to their aSI-H counterparts (**Fig. 4c**, **Supplementary Table 8b,c**).

**Fig. 4.**
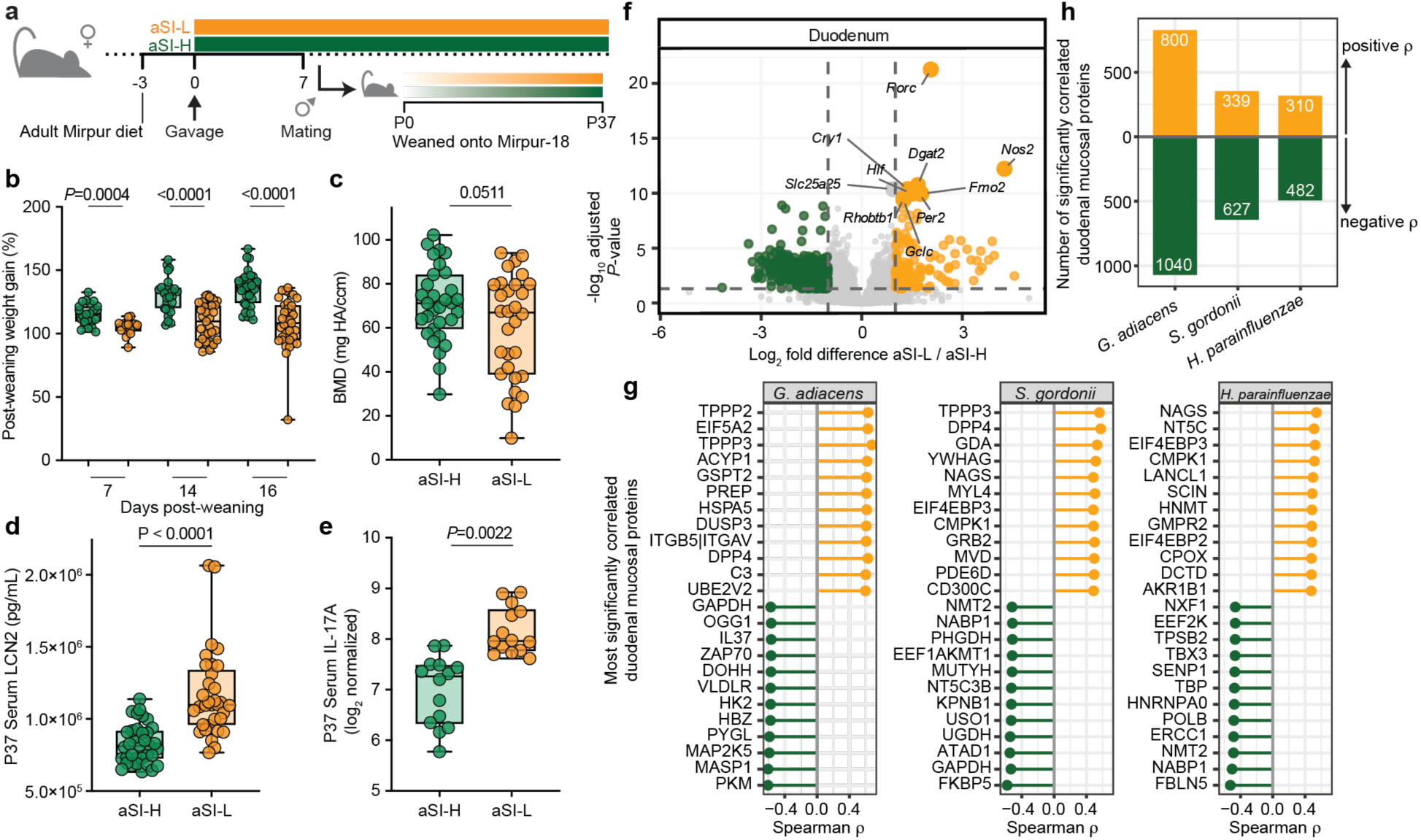
Effects of intergenerational transmission of aSI-L and aSI-H consortia in P37 offspring. **a,** Experimental design; numbers denote experimental day relative to timing of gavage. **b,** Serum LCN2 levels in P37 offspring (n=33-35 animals/group). **c,** Serum IL-17A levels in P37 offspring (n=14 mice/group). **d,** Femur bone mineral density in P37 offspring (n=30-31 animals/group). **e,** Pup post-weaning weight gain (n=35 animals/group; pups were weaned at P21). For panels (**b**-**e**), each dot represents an individual animal. **f,** Significantly differentially expressed genes in the duodenum of P37 aSI-L (positive, orange) compared to P37 aSI-H (green, negative) offspring (*DESeq2* Wald test, n=16 animals/group). **g-h,** aSI-L bacteria transmitted to P37 offspring that were identified in the duodenal aspirates of women and correlated with proteins in their duodenal biopsies. **g,** The 12 duodenal mucosal proteins in women that were most significantly positively and negatively correlated with the absolute abundances of *G. adiacens, S. gordonii* and *H. parainfluenzae* in aspirates (Spearman correlations, Benjamini Hochberg-adjusted *P*-values, n=78 women; see **Supplementary Table 9a**). **h,** The total number of significant positive and negative correlations between the absolute abundances of *G. adiacens, H. parainfluenzae* and *S. gordonii* in duodenal aspirates and protein levels in the duodenal mucosa (Spearman, Benjamini Hochberg-adjusted *P*-values, n=78 women).

Offspring of dams harboring the aSI-L consortium exhibited elevated systemic inflammation, as quantified by serum LCN2 (**Fig. 4d**), IL-17A (**Fig. 4e**) and MMP8 (*14*) (**Extended Data Fig. 7a**, **Supplementary Table 8a**). We subsequently quantified 96 serum proteins (O-link Target 96) in a subset of P37 offspring (n=14 mice/group). The cytokine CCL20 (C-C motif ligand 20, also known as Macrophage inflammatory protein-3a), which recruits immune cells, including Th17 cells, to the mucosa, was elevated in aSI-L offspring (**Extended Data Fig. 7b)**. Three proteins whose levels were significantly different between P37 aSI-L and aSI-H offspring were also significantly different in the plasma of undernourished compared to healthy women: CXCL1 (diminished in aSI-L and EED, **Extended Data Fig. 7c**), ENO2 (enolase 2) and IL-17 (both higher in aSI-L and EED, **Extended Data Fig. 7d**, **Supplementary Table 8a**).

### Intestinal transcriptional responses in P37 offspring

We performed bulk RNA-Seq on the duodenum and ileum of a subset of P37 offspring (n=15-16 mice/group). Genes that were more highly expressed in the duodenums of aSI-L compared to aSI-H offspring included (i) those involved in innate immunity (e.g., *Nos2*, *Duox2* and *Duoxa2*) (**Extended Data Fig. 7d,e**, **Supplementary Table 9d**), (ii) *Rorc*, the transcription factor that enables differentiation of Th17 cells (**Fig. 4f, Supplementary Table 8d**), in alignment with the elevated number of Th17 cells observed in aSI-L adult female mice (**Fig. 3d**), (iii) genes comprising Reactome pathways involved in antimicrobial responses (‘Alpha-defensins’) and (iv) genes related to DNA damage and repair (‘DNA Repair’, ‘DNA Double Strand Break Response’, ‘DNA Damage/Telomere Stress Induced Senescence’) (**Supplementary Table 8e**).

Genes whose expression was significantly elevated in the intestines in aSI-L P37 mice that are homologues of protein products enriched in the duodenal mucosal proteomes of women with EED include those involved in myeloid cell survival and proliferation (*Il34, Ly6d*), cellular responses to inflammation (*Fkbp5, Il4i1*), endothelial cell survival and vascular morphogenesis (*Kdr* which encodes VEGFR2, *Cd34*, *Mmp17*) (**Extended Data Fig. 8a,b**, **Supplementary Table 8f**).

### Taxa associated with barrier disruption and inflammation in women with EED and in gnotobiotic mice

We matched taxa whose absolute abundances were significantly different between aSI-L and aSI-H female mice and/or P37 offspring (**Supplementary Table 8g**) to taxa in the duodenal aspirates of the human donors (**Extended Data Fig. 9a**). We then correlated the absolute abundances of these taxa in the human duodenal aspirates with levels of proteins in the corresponding duodenal mucosal biopsies (**Fig. 4g**, **Supplementary Table 9a**). The results disclosed that the absolute abundance of *G. adiacens* had the greatest number of significant correlations with levels of duodenal mucosal proteins, followed by *S. gordonii* and *H. parainfluenzae* (**Fig. 4h**). These three bacteria were positively correlated with complement proteins (C3, C1QTNF5, C1QL3, C4A/B, C5/6), cytokines (IL-6R, IL-9, IL-15, IL-17RB, IL-18, IL-20RA, IL-21R, IL-25, IL-26), interferons (IFNA1, IFNA8, IFNGR1, IFNGR2), innate immune system modulators (ACE/ACE2 and HLA-G), pattern recognition receptors (SIGLEC1, SIGLEC6, SIGLEC14), and proteins involved in oxidative stress responses (AKR1B1, AKR1B10) (**Supplementary Table 9a**, **Extended Data Fig. 9b**). Conversely, these bacteria were significantly negatively correlated with levels of duodenal mucosal proteins involved in the Reactome signaling pathways for Insulin/IGF, Ras, VEGF, PDGF, FGF and GRH-R (Gonadotropin-releasing hormone receptor) in the human donors (**Extended Data Fig. 9c, Supplementary Table 9a,b**). Together, these data indicate that *G. adiacens, S. gordonii* and *H. parainfluenzae* are associated with pro-inflammatory immune responses and impaired intestinal barrier function in both gnotobiotic mice and women with histopathologically confirmed EED.

### Duodenal bacteria and mucosal proteins that characterize EED in adult Bangladeshi women and children

We compared the duodenal mucosal proteomes of women and children with EED, seeking proteins that were consistently elevated in adult women and children with histopathologic evidence of severe EED (histopathology score=3) compared to those with normal histology (**Fig. 5a**, **Extended Data Fig. 9**). These 142 (out of 1,114) proteins included several immuno-inflammatory proteins; Interleukin-18 receptor 1 (IL18R1), Neutrophil-activating peptide 2 (PPBP), Lysozyme C (LYZ), Toll-like receptor 4 (TLR4), Eotaxin (CCL11), Immunoglobulin A (IGHA1/2), Endothelial monocyte-activating polypeptide 2 (AIMP1), C-C motif chemokines 25 and 28 (CCL25/28), S100 Calcium-Binding protein-A9 (S100A9), Hexokinase 1 (HK1), Tumor Necrosis Factor Ligand Superfamily Member 14 (TNFRSF14) Complement component 1Q binding protein (C1QBP) (**Supplementary Table 9c**) and Lectin, Mannose-binding 2 (LMAN2), which was also elevated in plasma of women with EED (**Supplementary Table 1d**). These results highlight the overlapping duodenal mucosal proteomic signatures of EED in women and children that may be assessed non-invasively.

**Fig. 5.**
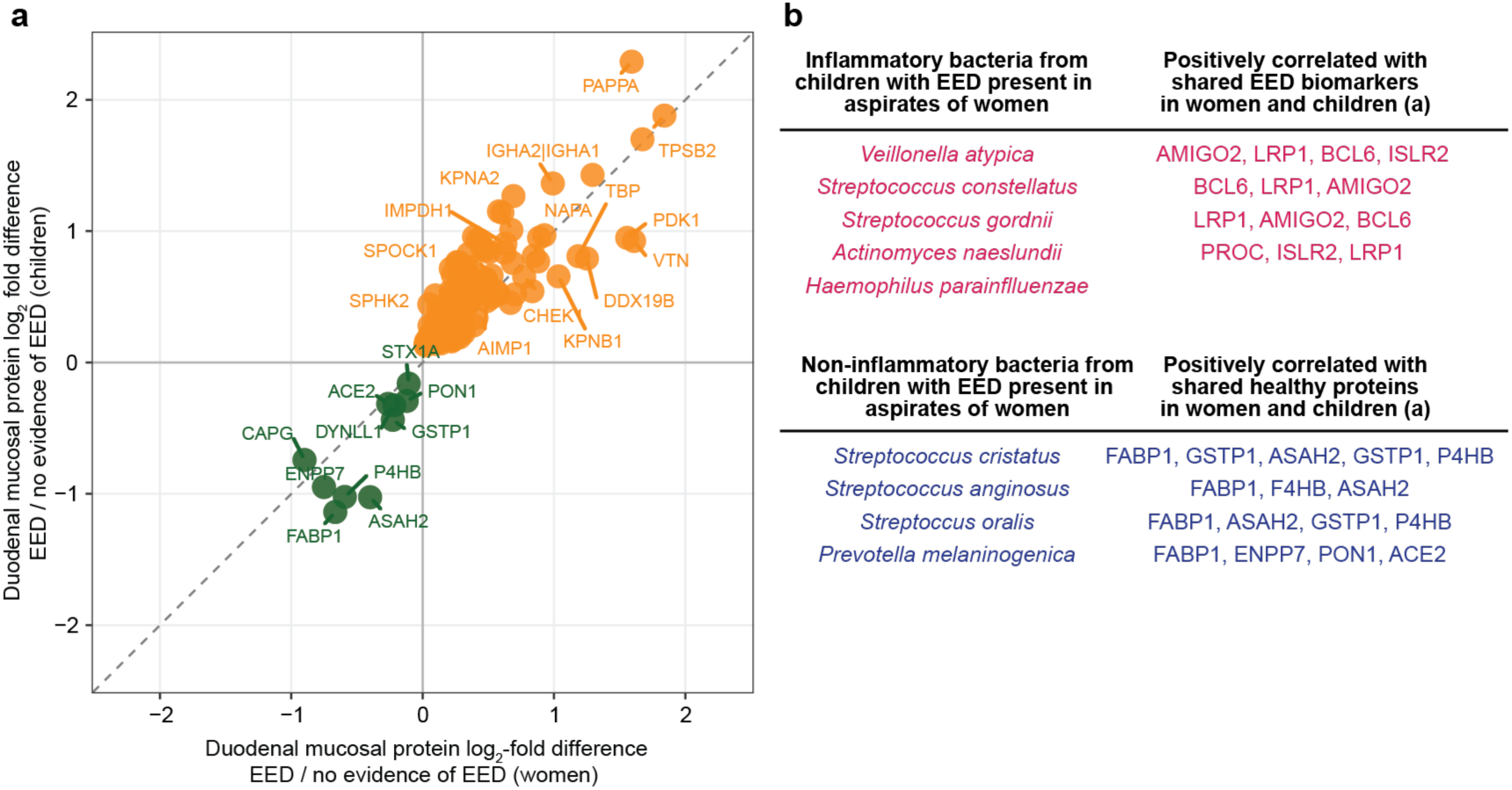
Duodenal mucosal proteins and bacterial taxa indicative of EED in women as well as children. **a,** Concordance between duodenal mucosal proteins whose levels were significantly higher (positive, orange) or lower (negative, green) in women and children with severe EED (histopathology score 3; n=15/78 biopsies from women in the current study, 37/89 undernourished children in the BEED study^14^ whose LAZ failed to improve after nutritional intervention) compared to those without histopathologic evidence of EED (histopathology score 0; n=20/78 women in the current study, 5 out of the 89 children^14^, *limma eBayes*). **b,** Duodenal bacteria cultured from children with EED that were associated with elevated inflammation and impaired post-natal development (top, cSI-I) or healthy development (bottom, cSI-N) in a gnotobiotic mouse model of intergenerational transmission^16^. Proteins highlighted include (i) shared biomarkers of EED (elevated in women and children with EED in **a**) that were significantly positively correlated with the absolute abundances of cSI-I taxa in the aspirates of adult women (top) and (ii) shared ‘healthy’ proteins (diminished in women and children with EED compared in **a**) that were significantly correlated with the absolute abundances of cSI-N taxa.

We recently described an analogous preclinical model where a consortium of duodenal bacteria that had been cultured from stunted Bangladeshi children with EED was introduced into germ-free female mice with subsequent dam-to pup transfer; the recipient pups exhibited impaired growth and elevated intestinal inflammation. The consortium that produced these phenotypes was named ‘child small intestinal inflammatory’ cSI-I^16^, **Supplementary Table 9d**). Another consortium assembled from these children that did not produce this inflammatory response upon dam-to-pup transmission served as a reference control (child small intestinal non-inflammatory consortium, abbreviated cSI-N^16^).

We found that members of the cSI-I consortium were also present in aspirates of adult women described in this study – they included *Streptococcus constellatus, Veillonella atypica, Actinomyces naeslundii, H. parainfluenzae,* and *S. gordonii* (**Fig. 5b**). In adult women, the absolute abundance of *S. gordonii* increased with severity of EED (**Fig. 2a**). Furthermore, the absolute abundances of these five taxa were significantly positively correlated with immunoinflammatory proteins in the duodenal mucosa, including those involved in migration and activation of immune cells (e.g., components of pathways for ‘Regulation of T cell activation’, ‘Regulation of macrophage migration’, ‘Regulation of leukocyte differentiation’), complement activations, and ‘Detoxification of Reactive Oxygen Species’ (**Supplementary Table 9e,f**).

In contrast, duodenal bacteria from the ‘child small intestinal non-inflammatory’ consortium included (i) *Prevotella melanogenica, Streptococcus anginosus,* and *Streptococcus cristatus* (**Fig. 5b**) – these species trended towards increased absolute abundance in the aspirates of women lacking histopathologic evidence of EED (see **Supplementary Table 2c** for statistics), and (ii) *Streptococcus oralis*, whose abundance was significantly negatively associated with histopathology score (**Fig. 2a**, **Supplementary Table 2b**). Moreover, the absolute abundances of *S. cristatus, S. anginosus,* and *P. melanogenica* in duodenal aspirates of adult women were significantly negatively correlated with proteins involved in innate immune signaling pathways (i.e., when the abundances of these bacteria are higher, expression of these innate immune signaling pathways decreases, **Supplementary Table 9e,g**). *P. melanogenica* and *S. cristatus* are reported normal inhabitants of the oral cavity^32,33^.

Together, these results indicate conserved strains within the small intestinal microbiota that contribute to shared pathogenic pathways underlying EED in both adult Bangladeshi women and children living in the same urban locale. These taxa were transmitted from dams to pups in our intergenerational gnotobiotic mouse models where they contributed to pathology in offspring.

## DISCUSSION

Maternal undernutrition is a key predictor of undernutrition in infants and children^6^; however, EED in adult women of reproductive age has been understudied. Our previous work demonstrated that bacteria in duodenal aspirates from stunted children with EED correlated with markers of enteropathy and growth impairment^14^. Here, we extend this framework of defining associations between members of the duodenal microbiota with intestinal tissue and plasma proteins to Bangladeshi women of reproductive age. Importantly, inclusion of well-nourished women with normal BMI from the same locale as those with EED in our esophagogastroduodenoscopy (EGD) study advances our understanding of the mechanisms underlying EED since comparable ‘healthy’ controls were not available in our prior studies of children. Furthermore, focusing on expressed proteins complements prior mRNA transcript-focused investigations that also lacked geographic location-matched healthy reference groups^34,35^. By comparing associations between duodenal and fecal bacteria with the duodenal mucosal and plasma proteomes of women with and without histopathologic evidence of EED, we identified a set of candidate plasma protein biomarkers of immunoinflammatory responses observed in the duodenal mucosa of nonpregnant women with EED. This set of plasma biomarkers, identified in this Bangladeshi study population, can serve a starting point for their measurement, using a variety of assay formats, in other sites in the global South (and elsewhere) where the burden of maternal undernutrition is great.

We determined whether discordant features of the duodenal microbiota were causally related to enteropathy by using gnotobiotic mice colonized with bacterial consortia derived from undernourished compared to well-nourished women. Proteomic, snRNA-Seq, and flow cytometric analyses demonstrated that an EED-associated, inflammation-inducing consortium induced intestinal barrier dysfunction that was linked to a Th17 mucosal immune response in recipient adult gnotobiotic female mice. Moreover, dam-to-pup transmission of the inflammatory consortium produced enteropathy in offspring, accompanied by significant reductions in weight gain. Bacterial species whose abundances were elevated in the intestines of offspring of dams harboring the inflammatory consortium were also significantly correlated with duodenal mucosal proteins that were discriminatory for EED in Bangladeshi women. The intergenerational transmission of these phenotypes in our model supports the hypothesis that vertical microbiota transfer is a mechanism that contributes to perpetuating the cycle of undernutrition and EED.

While enumeration of bacteria in the distal gut microbiome through sequencing analysis of fecal samples has been the standard approach in most studies of childhood undernutrition and EED^11,36–39^, the small intestine (SI) is the primary site of EED. Therefore, to focus on the role of the SI duodenal microbiota in EED, our mouse experiments did not include bacteria recovered from fecal samples; future studies are needed to ascertain whether there are modifying effects of reconstituting more complete intestinal communities using both SI and fecal communities from EED donors to colonize mice.

Longitudinal sampling in clinical trials is the gold standard; however, the invasive nature of EGD precluded our ability to obtain multiple luminal aspirates from the same individuals. Furthermore, less than 10% of the diversity in the duodenal microbiota of women was captured through our *in vitro* culturing and colonization of gnotobiotic mice, reflecting current technical limitations in modeling the diversity of human microbiota in controlled preclinical experiments. Future clinical trials using capsule-based technology^40^ could allow enhanced biogeographic sampling over time and expand the number of culturable organisms. In addition, decompartmentalization of oral bacteria to the small intestine^16,38,41^ and their subsequent contribution to pathophysiology of EED warrant further clinical investigation. Future studies comparing the oral and small intestinal microbiota of undernourished women with EED and their well-nourished counterparts without evidence of enteropathy prior to, during, and after pregnancy could provide insights into (i) the taxonomic and encoded functional features (as well as the expressed functions) of microbial communities occupying these different habitats, (ii) the relationships with these community features maternal health and offspring development, and (iii) potential microbial targets for therapeutic intervention in women and children. These therapeutic targets and candidate biomarkers/mediators of therapeutic responses can be tested in the type of gnotobiotic models we describe here and elsewhere^16^.

In summary, our study provides preclinical evidence that the small intestinal microbiota of women with EED contributes to their nutritional status and that maternal transmission of EED associated bacteria in this model has adverse consequences for offspring growth and development. The results suggest that improving gut health via microbiota-directed therapies may be one way to break the intergenerational cycle of undernutrition.

## MATERIALS AND METHODS

### Study Design

This study enrolled undernourished Bangladeshi women of child-bearing age who failed a standard nutritional intervention (one egg and 150 mL whole milk daily for 6 days/week for 2 months^15^) and their well-nourished counterparts so that esophagogastroduodenoscopy (EGD) could be performed with collection of duodenal aspirates and duodenal mucosal biopsies. Blood and fecal samples were collected concurrently. Bacteria were cultured from the duodenal aspirates of both undernourished women and well-nourished controls to evaluate their effects in gnotobiotic adult female mice and their offspring.

## HUMAN STUDIES

### Ethics and Inclusion

This work was performed as part of a long-standing collaboration investigating the role of the gut microbiota in undernutrition that is covered by a memorandum of understanding, between teams led by Tahmeed Ahmed (International Centre for Diarrhoeal Disease Research, (icddr,b) Bangladesh) and Jeffrey Gordon (Washington University in St Louis, USA). Bacterial isolates used in this study were obtained from duodenal aspirates collected from the previously reported Bangladesh EED study, which was approved by the Ethical Review Committee (ERC) at the icddr,b (protocol no: PR-16007; ClinicalTrials.gov number, NCT02812615). Written, informed consent was obtained from all participants. Human biospecimens were transferred to Washington University under a Materials Transfer Agreement (MTA). Bacterial isolates cultured and used in this study are the property of icddr,b and are available under MTA upon request to T.A. and J.I.G.

### Design and histopathologic scoring of duodenal mucosal biopsies

All biospecimens from the women in described in this report were collected with approval by the Ethical Review Committee at icddr,b. All participants provided informed, written consent. Details of study design, recruitment, and exclusion criteria for the clinical trials included in this study have been reported elsewhere^15,17,18^. Briefly, undernourished (BMI < 18.5 kg/m^2^) women 18-45 years old were recruited to the Bangladeshi EED (BEED) study for nutritional therapy. Exclusion criteria included severe anemia (Hb < 8 g/dL), chronic or acute diseases, pregnancy, lactation, drug abuse, known psychiatric disorders, and conditions causing secondary malnutrition. Women whose BMI failed to improve (<10% BMI increase) after three months underwent EGD for collection of duodenal biopsies and dry aspirates. Fecal samples and plasma were collected concurrently.

As part of the BEED study, well-nourished (BMI = 18.5-30 kg/m^2^) women 18-45 years old seeking treatment for dyspepsia who were residing in the same urban slum (Mirpur) in Dhaka, Bangladesh underwent EGD. Those who had normal endoscopic findings were included as healthy controls^15,17^, and duodenal biopsies and aspirates were collected. In the follow-up ‘Normal BMI’ cohort^18^, adult women from non-slum sections of Dhaka were included if they were 18-45 years of age and had BMI 20-25 kg/m^2^; duodenal biopsies and aspirates, plasma, and fecal specimens were collected.

Biopsy samples were fixed in 10% buffered formalin solution, and paraffin sections were prepared with hematoxylin and eosin (H&E) staining. Histological scoring criteria are described in detail in ref.^17^. Intestinal morphometric parameters measured included: villous atrophy, crypt hyperplasia, and inflammatory infiltrates in the lamina propria. Histological parameters were evaluated as follows: (i) villous atrophy – mild = mild reduction in villous height, subtotal = partial blunting, total = complete atrophy or flattening of villi; (ii) crypt hyperplasia – absent = no crypt elongation, present = crypt elongation or hyperplasia evident; (iii) inflammatory infiltrates – mild = mild increase of inflammatory infiltrates, moderate = lymphoid aggregates present, marked = diffuse and intense inflammatory infiltrates. These descriptive morphometric features were aggregated into histological scores from zero to three, with three indicating the most severe evidence of EED.

### Plasma and duodenal mucosal proteomics

#### Plasma

Blood (4.5 mL) was collected from undernourished (n=22) and normal-BMI women (n=25) in EDTA-coated tubes and was centrifuged at 3000 x *g* for 10 minutes (20 °C) to separate plasma, which was subsequently aliquoted and stored at −80 °C. A 55 µL aliquot of plasma was used to measure 9,609 unique proteins (SomaScan 11K Assay; SomaLogic).

#### Duodenal and plasma proteomic analyses

Duodenal biopsies were stored at −80 °C until extraction, at which time they were transferred to ice-cold Tissue Protein Extraction Reagent (T-PER, ThermoFisher 78510) supplemented with protease inhibitor (Roche cOmplete Ultra Protease inhibitor, 481761). Proteins were extracted with Cytiva sample grinding kits (80-6483-37) according to the manufacturer’s recommendations. Briefly, grinding tubes were centrifuged for 1 minute at 18,000 x *g* and excess liquid was removed. 200 µL of ice-cold T-PER containing protease inhibitor was aliquoted into each tube, which was maintained on wet ice. Biopsy samples were transferred to the tissue grinding tubes with sterilized fine point tweezers. Tissue was homogenized manually using the pestles provided in the Cytiva kit until no visible pieces of tissue remained. Homogenized samples were centrifuged at 18,000 x *g* at 4 °C for 20 minutes. The supernatant was transferred to a new 2 mL screw cap tube (Axygen). Tissue homogenates were diluted 1:100 into 1X PBS containing protease inhibitor (Roche cOmplete Ultra) and total protein was quantified using the Micro BCA Protein Assay kit (ThermoFisher 23235). Samples were normalized to 200 µg/mL in 1X PBS and stored at −80 °C. The SomaScan 7K Assay (SomaLogic) was used to measure 6,401 unique SOMAmers (which bind specific proteins and serve as a proxy for the abundance of the corresponding protein) in duodenal biopsy homogenates.

Output from SomaScan proteomic assays were quality-checked and normalized according to the manufacturer’s recommendations using hybridization controls, followed by median signal normalization across pooled calibrator replicates within the run. Median signal normalization was performed using Adaptive Normalization by Maximum Likelihood (ANML). ANML-normalized data were log_10_-transformed. Any SOMAmers flagged as not passing pooled QC thresholds for determining the accuracy of median replicate signal compared to a reference were removed (one duodenal tissue biopsy sample was removed based; no plasma samples were removed).

Plasma and duodenal mucosal proteins with the lowest 10% expression were removed, resulting in 9,448 plasma and 6,626 duodenal tissue aptamers tested. Differential abundance analysis was performed using the R package *limma* (v3.60.6^42^). A linear model was used to fit each aptamer using *lmFit* with a design matrix specifying cohort as the coefficient for plasma samples and histopathology score as the coefficient to be estimated for duodenal biopsies. Empirical Bayes moderation of variance^43^ was applied with *eBayes* and Holm-Bonferroni multiple hypothesis correction was included with *topTable(p.adjust = ‘holm’)*.

Proteins identified in plasma samples were tested for their association with BMI using the Significance Analysis of Microarrays (SAM^44^). The same filtering criteria were applied to the plasma SomaScan data as for the differential protein abundance analysis. Using the R package *SAM* (built on *samr,* v3.0), a d-statistic was computed for ‘Quantitative’ response type, standard regression method, 10 k-nearest neighbors and 1000 permutations. The 90^th^ percentile FDR < 0.05 was used to select the delta value d=1.08; features with *q*-value < 0.05 were considered significant.

Enrichment analysis of plasma and duodenal mucosal proteins was performed using *clusterProfiler*^45^ (v4.12.6). The association of plasma aptamers with BMI (d-score) and duodenal mucosal aptamer differences by histopathology score (*limma* logFC) were used to rank proteins. Gene Ontology (GO) Biological Process, Molecular Function, and Cellular Component databases were queried with *gseGO(geneList, orgDb=org.Hs.eg.db, pAdjustMethod = ‘BH’, minGSSize=20, pvalueCutoff=0.05)*. Reactome pathway enrichment was analyzed with *gsePathway(geneList, pvalueCutoff = 0.1, organism= ‘human’, pAdjustMethod = ‘BH’, by=‘fgsea’). P*-values were adjusted using the Benjamini-Hochberg method.

### Sequencing full-length bacterial 16S rRNA amplicons generated from duodenal aspirate samples

#### DNA isolation from duodenal aspirates

Duodenal aspirates were thawed on wet ice anaerobically for concurrent culturing (see *‘Culturing bacteria from duodenal aspirates’*) and low biomass DNA extraction with the Arcturus PicoPure DNA Extraction Kit (KIT0103, Applied Biosystems). For duodenal aspirates obtained from the BEED study, a spike-in consisting of 4.95 x 10^5^ cells of *Alicyclobacillus acidophilus*^46^ was added. For duodenal aspirates obtained from the second healthy cohort, a premixed *Allobacillus halotolerans* and *Imtechella halotolerans* spike-in standard (ZymoBIOMICS D6320-10) was diluted in autoclaved 1X PBS to the same number of cells added to the samples from the BEED study (4.95 x 10^5^ of each strain) in a final volume of 30 µL. 50 µL of duodenal aspirate was added to the spike-in. Aspirates were centrifuged at 5,000 x *g* for 10 minutes at 10 °C. During centrifugation, lyophilized Proteinase K was centrifuged briefly prior to resuspension in 155 µL Reconstitution Buffer, then maintained on wet ice. After centrifugation of the aspirates, 50 µL of the supernatant was removed and 50 µL of reconstituted Proteinase K was added to the cell pellet with gentle pipetting to mix. Samples were incubated at 65 °C for 10 hours then 95 °C for 10 minutes in a digital oven (Quincy Lab, Inc Model 30E), prior to overnight freezing at −20 °C. 250 µL of 0.1 mm zirconia silica beads and one 3.97 mm steel ball were used to bead beat samples in a Biopsec Minibeadbeater-96 for 4 minutes in 500 µL 2X buffer A (0.2 M NaCL, 0.2 M Tris, 0.02 M EDTA), 210 µL 20% SDS, and 510 µL phenol:chloroform:isoamyl-alcohol (pH 8). Samples were centrifuged at 3,200 x *g* for 4 minutes at 4 °C; 180 µL of the resulting aqueous phase extract was then added to a 720 µL 675:45 mixture of Qiagen Buffer PM and 3 M NaOAc (pH 5.5), mixed, and subsequently transferred to a Qiagen QiaQuick Gel Extraction column. The remaining DNA purification steps were performed with Qiagen Buffer PE and Buffer EB according to the manufacturer’s specifications.

#### Library preparation, sequencing, and analyses

DNA concentration was quantified with Qubit. Kinnex full-length 16S rRNA libraries were prepared according to the manufacturer’s specifications. De-concatenation was performed using Read Segmentation and resulting demultiplexed reads were trimmed (TrimGalore v0.6.6 –quality 20). Adaptors were removed from trimmed reads with Cutadapt (-g AGRGTTYGATYMTGGCTCAG… AAGTCGTAACAAGGTARCY --error-rate 0.1 –revcomp). Subsequent processing of 16S rRNA reads was performed with the *dada2*^47^ pipeline with minor modifications to the *filterAndTrim* step *(minLen=1000, maxLen=1600, maxN=0, rm.phix=TRUE)*. Error rate (*learnErrors*) learning was performed for each fastq file; dereplication (*deRepFastq*) and dada steps were performed with standard parameters. Taxonomic predictions were generated with *assignTaxonomy* using the GreenGenes (2024) database.

In *phyloseq* ^48^ (v1.48.0), bacterial reads from duodenal aspirates were normalized to spike-in counts to approximate absolute abundance in cecal contents. For all analyses, ASV abundances were collapsed into species-level abundances using *tax_glom(taxrank = “Species”)*. Bacterial species with a prevalence of <50% of the smallest group of participants involved in each comparison were removed. In total, 21,493 unique ASVs and 887 species were identified in duodenal aspirate samples; 127 species were present in at least 10 samples (chosen for 50% of n=20 women with histopathology score 0). Statistical analysis of differences in the absolute abundances of bacterial taxa was performed using a zero-inflated Gaussian mixture model with *metagenomeSeq*^49^. Briefly, counts were normalized using cumulative-sum scaling (*cumNormStatFast*). The statistical model *(∼histopathology_score + extraction_batch*) was used to determine significant associations with histopathology score as a numeric with *fitZig*. Linear models comparing bacterial abundances in duodenal aspirates were performed on log_10_-transformed abundances treating histopathology score as a factor *(histopathology_score_group + extraction_batch* and *∼disease_status + extraction_batch*).

### Sequencing full-length 16S rRNA amplicons from human fecal samples

DNA from frozen fecal samples was extracted as described previously^50^. Briefly, fecal samples were pulverized in liquid nitrogen, ∼50 mg of pulverized material was homogenized by bead-beating with 500 µL 0.1 mm zirconia silica beads in a solution of 500 µL phenol:chloroform:IAA (25:24:1), 210 µL 20% SDS and 500 µL buffer A followed by purification (QiaQuick, Qiagen) and storage in Tris-EDTA buffer.

Fecal DNA samples were normalized to a concentration of 0.4 ng/µL. Libraries were prepared and analyzed as described in ‘*Sequencing full-length 16S rRNA amplicons generated from duodenal samples*’ without normalizing reads to exogenous spike-in counts. 200 species were present in at least 9 samples (i.e., 50% of the 19 women with a histopathology score of 0). Differential abundance testing at the species level was performed using *metagenomeSeq* as described above.

#### Enteropathogen detection by multiplex quantitative PCR

DNA extracted from fecal samples obtained from undernourished women or healthy controls was quantified via Qubit then normalized to 2 ng/µL. Levels of 19 common bacterial, viral, and protozoal gastrointestinal pathogens were determined with a microfluidic digital PCR system (Fluidigm Juno). Previously described primers^21^ that target unique sequences within each pathogen were used; mean C_t_ values < 30 designated a positive result (i.e., presence). Pathogen prevalence is reported as presence/absence in **Supplementary Table 4c**.

### Associating duodenal bacteria with duodenal mucosal proteins

To identify bacteria with abundances that were associated with host protein levels independent of EED status, we fit a main effects linear model for each bacterium–protein pair using the R package *limma* with the design formula: *aptamer ∼ bacterial_abundance + EED_status*, where *aptamer* represents log_10_-transformed SomaScan values that were filtered to remove the 30% of aptamers with the lowest variance, *bacterial_abundance* represents log_10_-transformed absolute abundances in duodenal aspirates or relative abundances in fecal samples, and *EED_status* is a binary indicator of disease status (histopathology score ≥ 1 or histopathology score = 0). For each of the bacterial taxa, all protein targets were fit with the linear model using *lmFit*, and the ß_1_ coefficient was extracted (slope). Empirical Bayes moderation of residual variances was implemented with *eBayes(robust = TRUE)*. Multiple hypotheses were corrected for using the Benjamini-Hochberg false discovery rate correction across all bacterium-protein pairs. Associations with *q*<0.05 were considered significant. Bacteria were included if they were present in more than 20% of samples and where matched biopsies were available (n≥16 women).

To identify bacteria-protein pairs whose linear relationship differed significantly between women with and without EED^51,52^, we used the following design formula: *aptamer ∼ bacterial_abundance + EED_status + bacterial_abundance:EED_status*. This model yielded the main effect of bacterial abundance in healthy women (ß_1_), the effect of EED status (ß_2_) and the interaction term (ß_3_) representing the differences in slopes between bacterial and protein abundance. As above, empirical Bayes moderation and multiple hypothesis correction were applied. Enrichment analyses on proteins that were significantly associated with bacterial abundance in this disease-specific model were performed using the R package *gProfiler2*^53^ with the function *gost(query, organism = “hsapiens”, correction_method = “fdr”, significant = TRUE, evcodes = FALSE, mutli_query = FALSE)*. Proteins that were significantly linearly related to the abundance of bacteria either positively or negatively were queried separately. Bacteria were included if they were present in ≥ 9 samples from each group (i.e.,women with or without EED).

#### Dimensionality reduction of human bacterial and proteomic datasets

Dimensionality reduction of duodenal and fecal bacterial taxa was applied using the R package *mixOmics* (v6.28.0)^54^. Sparse partial least-squares discriminant (sPLS-DA)^20^ was used to predict taxa (ASVs) that were associated with cohort (fecal samples) or histopathology score (duodenal aspirates). Bacterial abundances were centered and log ratio-transformed using a tuning step with M-fold validation as follows: *tune.splsda(ncomp=6, logratio=‘CLR’, validation=‘Mfold’, folds=5, nrepeat=50).* The number of features (bacterial species) to test from each dataset was defined with *c(1:10, seq(20, nASVs, 10))* where *nASVs* was the total number of species included in each analysis. The final sPLS-DA model was run with *splsda(logratio=‘CLR’, ncomp, keepX)* where the number of components *ncomp* and number of features *keepX* were defined using the output of *tune.splsda*. The sPLS-DA models were evaluated with *perf(folds=5, validation=‘Mfold’, dist=‘max.dist’, nrepeat=50*).

A sPLS-DA model for duodenal mucosal proteins was fit using log_10_-transformed data that were normalized as described in ‘*Duodenal and plasma proteomic analyses*’, using biopsy histopathology score as the predictive variable *splsda(Histopathology_Score, ncomp=3)*.

#### Data Integration Analysis for Biomarker discovery using Latent cOmponents (‘block’ sparse partial least squares discriminant analysis)

Block sPLS-DA was used^55^ to integrate bacterial abundance in duodenal aspirates with duodenal mucosal proteomic data. SomaScan datasets were filtered to remove the 30% of aptamers with the lowest variance then centered (*scale(center=T)*). Bacterial abundances were log ratio-transformed and centered; the 10% of species with lowest variance were removed. Clinical cohort (undernourished vs. healthy) was used as the categorical variable for the fecal bacteria-plasma proteome comparison while histopathology score was used for the duodenal aspirate bacteria-duodenal mucosal proteome comparison. The final block sPLS-DA models were run with *block.plsda(ncomp=5, design, ncomp, keepX)*, where *design* was created using the correlation between the X and Y features resulting from *pls(ncomp=1)* on the bacterial abundance and SomaScan matrices. The models were evaluated with *perf(validation=‘Mfold’, folds=10, nrepeat=10)*, which was used to provide the optimal number of components, *ncomp.* The number of features (*test.keepX*) to include in the test was defined with *c(seq(10,250,25))* for bacteria and *c(seq(10,1000,100))* for aptamers. Block sPLS-DA models were tuned with *tune.block.splsda(ncomp, test.keepX, design, validation=‘Mfold’, folds=f, nrepeat=1, dist=‘centroids.dist’)*, where *f=8* for the duodenal bacteria-proteome model and *f=10* for the fecal bacteria-plasma proteome model. The tuning function provided the number of bacterial species and aptamers (features in *keepX*) to retain in the final model.

### Culturing bacteria from duodenal aspirates

The methods used for culturing bacterial strains from duodenal aspirates obtained from adult women enrolled in the BEED study were analogous to those we employed previously^14^. All culturing materials were pre-reduced in a Coy chamber, under an atmosphere of 75% N_2,_ 20% CO_2_ 5% H_2_, for at least 24 hours. Duodenal aspirates were thawed under anaerobic conditions on wet ice. A 100 µL aspirate sample was diluted 1:4 in 300 µL 1X PBS. 100 µL aliquots of diluted aspirate were plated on two Brain Heart Infusion plates supplemented with 10% defibrinated horse’s blood (BHI-BA) and two Chocolate Agar plates (Hardy Diagnostics, H25). One BHI-BA plate and one Chocolate Agar plate were incubated at 37 °C under anaerobic conditions. The other two plates were grown at 37 °C under microaerophilic conditions (NuAire In-VitroCell Direct Heat CO_2_ incubator, 85% N_2_, 10% CO_2_, 5% O_2_). Every 24 hours, colonies were picked into 600 µL of BHI broth in a 96-well plate containing 1 mL wells. After 48 hours growth at 37 °C under the same atmospheric conditions, strains were stored at −80 °C in 100 µL of a 1:1 mixture of 30% glycerol (prepared with 1X PBS).

#### Pooling

Strains were stored in 96 well plates at −80 °C in PBS containing 15% glycerol (v/v). For isolates originally isolated under anaerobic conditions, plates were thawed under anaerobic conditions and 20 μL aliquot of each stock was used to inoculate a well in a 1 mL deep-well plate (Thermo Scientific) containing 600 µL BHI broth (followed by anaerobic growth at 37 °C. For isolates originally isolated under microaerophilic conditions, a 20 µL aliquot of each stock was inoculated into 600 µL BHI broth prior to microaerophilic growth at 37 °C.

After incubation for 72 hours at 37 °C, a 20 μL aliquot of each anaerobic or microaerophilic culture was added to 600 µL fresh medium (sub-culture) which was incubated for an additional 48 hours at the same temperature under the same atmospheric conditions. Equal volumes of each isolate sub-culture were then pooled in 50 mL conical tubes anaerobically and the mixture was centrifuged at 2,500 x *g* for 15 minutes. Under anaerobic conditions (75% N_2,_ 20% CO_2_ 5% H_2_), the supernatant was removed, the pellet was resuspended in 30% glycerol, using 1/5 of the original culture volume and the pooed culture collections were aliquoted into 1.8-mL crimp glass vials (Wheaton), which were sealed and stored at −80 °C.

#### Strain identification

Individual bacterial isolates were identified by Sanger sequencing of full-length 16S rRNA amplicons generated by primers 8F and 1391R. An aliquot of each isolate’s broth culture that had been stored in PCR plates at −20 ℃ were thawed; 5 μL of that aliquot was transferred to a separate PCR plate and 45 μL ‘lysis buffer’ (25 mM NaOH, 0.2 mM EDTA) was added. The PCR plate was sealed with aluminum tape, incubated at 95 ℃ for 30 minutes then centrifuged briefly at 1,000 x g to gather condensate. 50 μL of ‘neutralization buffer’ (0.040M Tris-HCl) was added to the culture with lysis buffer. The plate was then centrifuged for 6 minutes at 3,000 x g. For each sample, 12.5 μL of One Taq Hot Start 2X (New England Biolabs M0484S), 0.5 μL of both 8F and 1391R primers (10 μM), and 9.5 μL of nuclease free water were added to 2 μL of lysed template DNA. PCR was performed using a Bio Rad C1000 Touch Thermal Cycler and the following cycling conditions: 30 seconds at 94 ℃ for the initial denaturation, followed by 34 cycles of 15 seconds at 94 ℃ (denaturation), 15 seconds at 52 ℃ (annealing), and 90 seconds at 68 ℃ (extension). This was followed by a 5-minute final extension at 68 ℃.

Successful generation of PCR amplicons was confirmed by gel electrophoresis using a 1.5% agarose gel in 1 x TAE buffer with 1 kb DNA ladder for reference. Samples with visible bands were sent to Azenta Life Science for enzymatic PCR clean up and Sanger sequencing. Raw sequences were trimmed and aligned with Geneious Prime (http://www.geneious.com). Trimmed reads were then assembled *de novo* using Geneious Prime’s ‘De Novo Assemble’ function. Taxonomic assignments were based on alignments obtained with BLAST utilizing the NCBI database. The best quality hit was selected based on the ‘Grade’ score given by the software; this score considers query coverage, e-value, and identity values. Only scores with a ‘Grade’ above 90% were considered. The isolate with the best quality hit for each unique species type was then selected for generation of permanent stocks.

## GNOTOBIOTIC MOUSE EXPERIMENTS

All experiments involving mice were performed using protocols approved by Washington University Animal Studies Committee. Mice were housed in plastic flexible film gnotobiotic isolators (Class Biologically Clean Ltd., Madison, WI) at 23 °C under a strict 12-hour light cycle (lights on at 0700h). Autoclaved paper ‘shepherd shacks’ were kept in each cage to facilitate the natural nesting behaviors and for environmental enrichment.

***Diets*** - The ‘Adult Mirpur’ diet was designed based on 24-hour dietary recall surveys and food frequency questionnaires taken from adults living in the Mirpur district of Dhaka, Bangladesh who were enrolled in the BEED study. A pelleted, sterile version of this diet was manufactured by Dyets, Inc. (Bethlehem, PA). The quantity of each ingredient used to prepare the diet is provided in ref.^16^. The composition of the ‘Mirpur-18’ diet was based on Bangladeshi complementary feeding practices for 18-month-old children living in Mirpur, as defined by quantitative 24-hour dietary recall surveys conducted in the MAL-ED study^56^. This diet was manufactured according to previously described protocol^14^.

The sterility of irradiated diets was confirmed by culture in (i) BHI broth, (ii) Nutrient broth, and (iii) Sabouraud-dextran broth (all from Difco) for one week at 37 °C under aerobic conditions, and in reduced Tryptic Soy broth (Difco) supplemented with 0.05% L-cysteine HCl under anaerobic conditions. All diets were stored at −20 °C prior to use. Nutritional analysis of each irradiated diet was conducted by Nestlé Purina Analytical Laboratories previously (St. Louis, MO), see ref.^16^.

### Generation of the aSI-L and aSI-H consortia

4-5-week-old germ-free C57Bl/6J mice were given *ad libitum* access to a standard chow diet (Diet 2018S, Envigo). Three days prior to colonization, mice were switched to the Adult Mirpur diet and were given *ad libitum* access throughout the course of the experiment. For initial characterizations of gnotobiotic mice orally gavaged (oral gavage needle; Cadence Science; catalog no. 7901) with the pooled Low BMI or Healthy BMI culture collections, animals were weighed and fecal samples were collected within gnotobiotic isolators. Animals were euthanized 18 days after colonization and LCN2 and MMP8 levels were assessed in serum (see *Protein measurements from serum and intestinal tissue*).

Flash-frozen cecal contents stored at −80 °C from mice with the highest and lowest levels of serum biomarkers of EED that were colonized with the pooled Low BMI or Healthy BMI culture collections were selected for clarification. Individual mouse cecal contents were weighed and homogenized in pre-reduced 30% glycerol prepared in 1X PBS and supplemented with 0.05% L-cysteine; homogenization was achieved with 2 mm glass beads under anoxic conditions in a 50 mL conical tube. After filtration through a 100 µM cell strainer (Corning) 30% glycerol was added to a final concentration of 1% w/v. These clarified cecal contents were aliquoted to 2 mL crimp top glass vials under anaerobic conditions prior to storage at −80 °C.

### Husbandry and tissue collection from adult females and offspring

#### Husbandry

6-8-week-old germ-free C57Bl/6J mice were given *ad libitum* access to an autoclaved breeder chow (Lab Diet 5021; Purina Mills, Richmond, IN) until 3 days prior to colonization, at which time they were switched to the Adult Mirpur diet for the remainder of the experiment. Mice received 200 µL of the stock solutions of the aSI-L or aSI-H consortia via an oral gavage needle.

One week after initial gavage, trio matings were performed (two females with one male). Pups born to these mothers were maintained with their dams in the same cage until weaning on postnatal day 21 (P21), at which time pups from the same litter were transferred to new cages and weaned onto the Mirpur-18 diet. For dams and their pups, bedding was replaced every 7 days and diets were provided *ad libitum*. Following weaning at P21, offspring weights were monitored and fecal samples were acquired.

#### Tissue collection from adult female mice

At the time of euthanasia, blood was collected retro-orbitally in serum separator microtainer tubes (BD) and kept at room temperature prior to centrifugation at 2,000 x *g* for 10 minutes; separated serum was aliquoted into 2 mL screw cap tubes and stored at −80 °C.

The intestine was removed and dissected on a disposable dissection board placed on wet ice. The following sections of the duodenum, jejunum, and ileum were flushed with PBS and pinned out in 10% neutral buffered formalin for histologic assessment: 3-5 cm from the gastroesophageal junction (GEJ), 15-17 cm from the GEJ, and 3-5 cm from the ileal-cecal valve (ICV). The following sections of the duodenum, jejunum, and ileum were cleared of intestinal contents with gentle extrusion with curved forceps then flash-frozen in liquid nitrogen and stored at −80 °C for snRNA-seq: 2-3 cm from the GEJ, 14-15 cm from the GEJ, and 1.5-3 cm from the ICV. Sections of the duodenum (1-2 cm from the GEJ) and ileum (6-7.5 cm from the ICV) were cleared of intestinal contents and flash frozen for protein assays.

#### Tissue collection from P37 offspring

Serum was collected as described above. The intestine was removed and dissected on a disposable dissection board placed on wet ice. The small intestine was divided into thirds (approximating the duodenum, jejunum, and ileum) and intestinal contents from each portion of the small intestine and colon were removed by gentle extrusion with curved forceps, weighed in 2 mL screw cap tubes, and snap-frozen in liquid nitrogen. Cecal contents were sampled using 10 µl plastic loops, frozen in liquid nitrogen then placed in 2 mL screw cap tubes. From the most proximal end of each portion of the small intestine, three 1 cm tissue segments were acquired, which, in addition to the remaining tissue segment, were stored in separate 2 mL screw cap tubes, flash-frozen in liquid nitrogen prior to storage at −80 °C. For the colon, two 1.5 cm segments were collected plus the remaining colonic tissue.

### FITC-4 kDa dextran gut permeability assay

After 4 hours of fasting, mice were gavaged with 80 mg/mL FITC-4 kDa dextran (Sigma Aldrich) at 0.6 mg/g. Food and water were withheld for an additional 4 hours, followed by collection of blood and euthanasia. Serum was isolated with a BD microtainer and fluorescence was measured using an excitation wavelength of 485 nm and emission wavelength of 530 nm with the Tecan Spark 20M plate reader.

### Micro-computed tomography (µCT) of bone

The femur was harvested from the left rear leg of P37 offspring and cleaned of muscle and connective tissue. Femurs were stored in 10 mL 10% neutral buffered formalin at room temperature for 24 hours. Fixed femurs were then washed in 1X PBS for 15 minutes; this wash step was repeated two more times, followed by a 30% ethanol wash for 30 minutes, a 50% ethanol wash for 30 minutes and a final 70% ethanol wash for 30 minutes. Femurs were subsequently embedded in 2% agarose and scanned with a μCT 40 desktop cone beam instrument (ScanCO Medical, Brüttisellen, Switzerland). For analyses of cortical bone, 100 slices were taken for each sample in the transverse plane, with a 6 µm voxel size (high resolution); slices began at the midpoint of the femur and extended toward the distal femur. The boundaries and thresholds for bone were drawn manually using μCT 40 software. Volumetric parameters were quantified using software associated with the ScanCO instrument.

### Histomorphometric and immunohistochemical analysis of intestinal tissue

Segments from the duodenum, jejunum, and ileum (see ‘Tissue collection from adult female mice’) were fixed in 10% neutral buffered formalin for 1 day, then washed and stored in 70% ethanol, all at 4°C. Tissue segments were positioned in agar then embedded in paraffin followed by preparation of 5 μm sections on slides that were stained with hematoxylin and eosin then scanned with a Hamamatsu NanoZoomer at 20X resolution for histomorphometric measurements of 10 well-oriented villus-crypt units/tissue segment/mouse using *QuPath*^57^. For immunostaining, additional 5 μm sections on slides were prepared and deparaffinized with successive immersion in xylene, ethanol, and water mixtures (*i.e.* 100% xylene, 50% xylene/50% ethanol, 100% ethanol, 95% ethanol, 70% ethanol, 50% ethanol, and water). Slides were then boiled in Tris-EDTA Buffer pH 9.0 buffer (Abcam, ab93684) for antigen retrieval, blocked with 10% normal goat serum (Abcam, ab7481) for 2 hours at room temperature, and incubated overnight at 4 °C with anti-ZO-1 primary antibody (1:100, ThermoFisher Scientific #61-7300) diluted in antibody diluent (Abcam, ab64211). Sections were subsequently washed with TBS-Triton-X 100 and incubated with goat anti-rabbit IgG–Alexa Fluor 647 (1:500, ThermoFisher Scientific #A-21245) for 2 hours. Stained sections were mounted with Fluoroshield Mounting Media containing DAPI (Abcam, ab104139) and scanned with a 3DHistech P250. Sections were analyzed in a blinded fashion using *QuPath*^57^ where villus peripheries were traced (3 villi/intestinal segment for n=4 mice/treatment group) and exported to *FIJI*^58^ for quantification of mean fluorescence intensities and normalization to background fluorescence intensities. Representative high-resolution images were collected with the Zeiss LSM 880 microscope (under 63X magnification/1.4 NA oil).

### Protein measurements in mouse serum and intestinal tissue

Serum that had been obtained at the time of euthanasia and flash-frozen upon separation with a BD microtainer, was thawed on ice and diluted 1:1000 for use with the Mouse Lipocalin-2/NGAL DuoSet ELISA kit (R&D Systems), 1:30,000 for use with the Mouse Complement C3 ELISA kit (Abcam), or subjected to the Mouse Cytokine/Chemokine 32-Plex Discovery Assay® Array (MD32) for measurement of IL-6 and IL-17A (Eve Technologies) or an O-link Target 96 Mouse Exploratory assay (Thermo Fisher; see **Supplementary Table 8a(ii)** for a list of proteins assayed). Frozen intestinal tissue was homogenized by (i) adding a 20-30 mg segment from the duodenum or ileum (see ‘Tissue collection from adult female mice’), to a vial with 1.6 mm aluminum oxide and silicon carbide particles (Matrix F; MP Biomedicals) and 600 µL cold T-PER buffer (Thermo Fisher Scientific) with Complete Ultra protease inhibitor (Roche) followed by (ii) bead beating for 1 minute then centrifugation at 13,000 x g for 5 minutes at 4 °C. Protein concentrations in the resulting supernatants were measured (Pierce BCA Protein Assay Kit; Thermo Fisher Scientific) and adjusted to 1 mg/mL using sterile PBS with protease inhibitor (Roche). IL-22 levels were measured with the Mouse Cytokine Th17 12-Plex Discovery Assay® Array (MDTH17-12, Eve Technologies).

### Flow cytometry of immune cell populations

Mice received a retro-orbital injection of 2 μg of anti-CD45 antibody to label intravascular leukocytes. Animals were euthanized 3 minutes post injection and small intestinal leukocytes were isolated as previously described^59^. Briefly, small intestines were flushed to remove luminal contents, and Peyer’s patches and mesenteric fat were removed. Tissues were opened longitudinally and incubated for 20 min with gentle agitation in Hanks’ balanced salt solution (HBSS) supplemented with 1 M HEPES (Corning), 10% bovine calf serum (HyClone), and 0.5 M EDTA (Corning) to remove epithelial cells, followed by vortexing. After a second round of gentle agitation and vortexing in fresh HBSS/bovine calf serum/EDTA buffer, the tissue was washed with HBSS and digested in Complete RPMI 1640 medium (Gibco) containing collagenase IV (Sigma–Aldrich) for 40 minutes at 37 °C with agitation. The resulting cell suspension was filtered through a 100-µm strainer (Falcon) and leukocytes were enriched by density gradient centrifugation using 40% and 70% Percoll (GE Healthcare). Cell suspensions were incubated with Fc block (clone 2.4G2, made in-house from HB-I97 hybridoma cells), treated with a viability dye (Live/Dead Aqua, Thermo Fisher Scientific), then subjected to staining for surface markers. The Foxp3 Staining buffer set (eBioscience) was then applied, followed by staining of intracellular proteins. The antibodies used are listed in **Supplementary Table 7g**. Samples were analyzed using the BD FACSymphony A3 flow cytometer and *FlowJo* (Treestar); gating strategies are shown in ***Supplementary Results***.

### snRNA-Seq of duodenum, jejunum and ileum

#### Isolation of nuclei and library preparation

Intestinal segments from the duodenum, jejunum, and terminal ileum (see *‘Tissue collection from adult female mice’*) were processed as previously described^60^. Tissues were minced in a buffer of 25 mM citric acid, 0.25 M sucrose, 0.1% NP-40, and 1X protease inhibitor (Roche); sheared using a Dounce homogenizer (Wheaton) to release nuclei; washed 3 times with a buffer of 25 mM citric acid, 0.25 M sucrose, and 1X protease inhibitor (Roche cOmplete Ultra); filtered through 100 μm-, 40 μm-, 20 μm-, and 5 μm-diameter strainers, and resuspended in a buffer consisting of 5 mM KCl, 3 mM MgCl_2_, 50 mM Tris, 1 mM DTT, 0.4 U/μL RNase inhibitor (Sigma), plus 0.4 U/μL Superase inhibitor (ThermoFisher Scientific). The Chromium Next GEM 3’ kit v3.1 (10x Genomics) was used to generate gel bead-in-emulsions then cDNA libraries using 7000 nuclei/sample. After quality assessment with a BioAnalyzer, balanced libraries were sequenced on an Illumina NovaSeq 6000 instrument [150 nt paired-end reads; 2.74 x 10^8^ ± 3.35 x 10^7^ reads/sample (mean ± SD)].

#### Data analysis

Sequencing reads were demultiplexed and aligned to the mouse reference genome (GRCm38/mm10) with *CellRanger* 5.0 (10x Genomics) and denoised using *CellBender*^61^. Sample datasets were further processed using the Seurat v4 framework^62^ to remove low quality nuclei (< 400 UMI, < 200 or > 8000 genes, > 5% mitochondrial reads, and/or > 60% ribosomal reads), normalize counts (*SCTransform*), remove predicted doublets (*DoubletFinder*), integrate samples within each tissue type (duodenum, jejunum, or ileum) using anchors identified by reverse PCA, and to cluster nuclei using *FindNeighbors* (dimensions 1:30) and *FindClusters* (resolution 1.4). Cell types were annotated by matching cluster gene markers (*FindMarkers*) with reported cell-type markers^26,27,63^. ‘Pseudobulk’ analysis was performed by aggregating counts within each cell type designation for clusters with > 100 nuclei/group, then performing comparisons across treatment groups with *DESeq2*. Gene set enrichment analysis (GSEA) was performed with *fgsea*. Human homologues to the mouse genes that were differentially expressed in the duodenums of aSI-L and aSI-H adult female mice were identified using the R package *homologene* with the function *human2mouse(db = homologeneData)*.

### Bulk RNA-Seq of duodenum, ileum and colon from P37 mice

#### RNA extraction and library preparation

RNA was extracted from 1-1.5 cm flash-frozen segments of the duodenum, ileum and/or colon (piece #1, *see “Tissue collection from P37 offspring” above*) using the Qiagen RNeasy 96 Kit. Total RNA was quantified (Qubit) and quality was assessed using a TapeStation (Agilent). cDNA libraries were generated using the Illumina Total RNA Prep with Ribo-Zero kit (Illumina). Barcoded libraries were sequenced on Illumina NovaSeq X Plus instrument [150 nt paired-end reads to a depth of 1.2×10^7^ ± 2.2×10^6^ reads/sample (mean ± s.d.).

#### Data analysis

Read quality was verified with *FastQC* (v0.11.7; http://www.bioinformatics.babraham.ac.uk/projects/fastqc/). Reads were trimmed to remove adapters and low-quality read segments using *trimgalore* (*trimgalore –quality 20 –fastqc*, v0.6.6; https://github.com/FelixKrueger/TrimGalore). Trimmed reads were pseudoaligned (*kallisto quant -b 100 –plaintext –bias –rf-stranded –pseudobam,* v0.48.0) to a *kallisto*^64^ index built from the Gencode v37 *Mus musculus* reference genome (*kallisto index gencode.vM37.transcripts.fa.gz*). MultiQC^65^ (v1.13) was performed on trimmed and mapped reads. Transcript counts were aggregated to gene counts. *Kallisto* ‘estimated counts’ values and transcripts per million (TPM) values were used to generate a gene-level expression matrix in R using *tximport*^66^ (v1.32.0) and *biomaRt*^67^ (v2.60.1). Gene annotations were acquired with *tximeta* [*tx2gene = tximeta(coldata, type = “kallisto’, cleanDuplicateTxps = T, useHub = T)*]. The ‘lengthscaledTPM’ function in *tximport* was used to adjust estimated counts by gene length and abundance [*tximport(x, type = ‘kallisto’, tx2gene, countsFromAbundance = ‘lengthScaledTPM’)*]. Raw counts were imported [*tximport(x, type = ‘kallisto’, tx2gene, countsFromAbundance = “no”)*] for *DESeq2*^68^ differential abundance analysis and filtered at a threshold of at least 10 counts across the minimum group size (n=16 animals). Differential expression on the resulting count matrix was performed for each tissue separately with *DESeq2* [*dds = DESeqDataSetFromMatrix(countData, colData*, *design = ∼Microbiota),* where ‘Microbiota’ denoted aSI-L versus aSI-H animals. *dds = DESeq(dds), res = results(dds, contrast = c(“Microbiota”, “aSI_L”, “aSI_H”)*].

EntrezIds were mapped onto gene IDs using *AnnotationDbi* (1.66.0; https://github.com/Bioconductor/AnnotationDbi) and *org.Mm.eg.db* (v3.19.1) [*mapIds(org.Mm.eg.db, keys = row.names(countData), keytype = “SYMBOL”, column = “ENTREZID”)*]. GSEA was performed using *clusterProfiler*^45^ (v4.12.6) querying the Gene Ontology (GO) [*gseGO(geneList, org.Mm.eg.db, ont = “ALL”, minGSSize = 20, nPermSimple = 10000, pvalueCutoff = 0.1, by = ‘fgsea’, eps = 0)*] and Reactome pathway databases [*gsePathway(geneList, organism = “mouse”, minGSSize = 10, nPermSimple = 10000, pvalueCutoff = 0.1, by = ‘fgsea’, eps = 0)*]. Human homologues to mouse genes that were differentially expressed in the duodenum, ileum and colon of P37 aSI-L and aSI-H offspring were identified using the R package *homologene* (https://github.com/cran/homologene) with the function *human2mouse(db = homologeneData)*.

### Quantitation of gut bacteria

#### DNA extraction

The absolute abundances of bacterial species were determined using methods described above with minor modifications. In brief, ‘spike-in’ bacterial strains whose genomes are easily differentiated from those of gut bacteria were added to each weighed frozen sample of intestinal contents prior to DNA isolation and preparation of barcoded libraries. For initial tests of the pooled culture collections and clarified cecal contents from mice harboring high and low levels of LCN2, *Alicyclobacillus acidiphilus* spike-in was added to frozen cecal contents after weighing (1.33 x 10^7^ cells per aliquot of cecal contents; DSM 14558; GenBank assembly accession: GCA_001544355.1). For experiments involving adult female mice and P37 aSI-L and aSI-H animals, a commercial 1:1 mix of *Imtechella halotolerans* and *Allobacillus halotolerans* was added (1.33 x 10^7^ cells per aliquot of cecal contents; ZymoBIOMICS product number D6320-10).

DNA was extracted from flash-frozen cecal contents obtained stored at −80 °C. To obtain crude DNA, frozen cecal contents (∼50 mg) were subjected to 4 minutes of bead-beating in a 2 mL tube containing 0.1 mm silica beads, one 3.97 mm steel ball, 500 µL phenol:chloroform:isoamyl alcohol and 710 µL of a 500:210 (v/v) mixture of 2X buffer A:20%SDS. Crude DNA was subsequently purified (QIAquick 96 PCR Purification Kit) and quantified (Qubit). The final DNA fragment size distribution was determined using an Agilent Technologies 4200 TapeStation. Purified DNA samples were normalized to 0.4 ng/µL prior to preparation of Kinnex PacBio full-length 16S rRNA amplicon libraries, as described above in ‘*Full-length 16S rRNA sequencing of human samples*’.

#### Differential abundance analyses

Raw PacBio HiFi reads were processed as described above in *Library preparation, sequencing, and analyses*. The resulting ASVs were collapsed to unique species using *tax_glom(taxrank = “Species”)*. Species counts were normalized to spike-in counts to approximate absolute abundance, as recommended by the manufacturer. Bacterial reads from mouse samples were aligned to the 16S rRNA sequences from the human datasets. Bacteria with <98.7% nucleotide sequence identity to a strain identified in humans were removed prior to statistical analyses, as well as bacteria that were detectable in fewer than 3 mice. The statistical significance of observed differences in the absolute abundances between groups was tested using a non-parametric Wilcoxon rank-sum test with FDR correction (Benjamini-Hochberg) on log_10_-transformed absolute abundances.

#### Matching taxa in mice with human samples

Best matches between bacterial taxa that colonized mice and their human donors were identified using two parallel approaches. For unique species present in gnotobiotic mice and duodenal aspirates or fecal samples, sequences were converted to *DNAStringSet* objects using the package *Biostrings* (v2.72.1; https://github.com/Bioconductor/Biostrings). First, a k-mer based approach was used to find the best matches between all mouse bacterial 16S rRNA sequences and all human duodenal aspirate or fecal bacterial 16S rRNA sequences using the package *stringdist* (v0.9.15) as follows: *stringdist(seq, ref_seq, method= ‘qgram’, q=12)*, the pairwise identities for the best matches were calculated with *pairwiseAlignment(seq1, seq2, type=‘global’).* Second, full alignments were used to compare the sequences of species-level representatives (from *tax_glom(taxrank=“Species”)*) that colonize mice with the sequences of species-level representatives in duodenal aspirates and fecal samples, again using *pairwiseAlignment*.

#### Correlations between taxa associated with pathology in mice and duodenal mucosal proteins in adult women

Taxa that were associated with pathology in P37 offspring were identified in the duodenal aspirates of women using the methods described above in ‘*Matching taxa in mice with human samples*’. Correlations between the abundances of these taxa and levels of duodenal mucosal proteins in matched samples were calculated using log_10_-transformed bacterial abundances and SomaScan data normalized as described above in ‘*Block sparse partial least-squares discriminant analysis*’. Spearman correlations [*cor.test(bacterial_abundance, protein_abundance, method=‘spearman’)*] were calculated for every possible bacterial-protein pair; *P*-values were adjusted using the Benjamini-Hochberg correction. Enrichment analyses were performed using the R package *gprofiler2*^53^ with the function *gost(query, organism = “hsapiens”, correction_method = “fdr”, significant = TRUE, evcodes = FALSE, mutli_query = FALSE)*, where the set of proteins that were significantly positively and negatively correlated with each bacterium were queried separately.

## Supporting information

Supplementary Results

## Acknowledgments

We thank David O’Donnell, Maria Karlsson, and the late Justin Serugo for their invaluable assistance with mouse husbandry, Martin Meier for generating libraries for shotgun sequencing and microbial RNA-Seq, and Jessica Hoisington Lopez, MariaLynn Crosby and members of the Genome Technology Access Core (GTAC) at Washington University School of Medicine for sequencing these libraries. We also thank Chris Sawyer, Andrew Lutz and Jinsheng Yu at GTAC for running and pre-processing data for the SomaScan assays reported in this manuscript. We are also grateful to Christopher Markovic for generation of and sequencing of Kinnex PacBio libraries, Chris Sawyer for assistance with the multiplex qPCR assay, Brooks Brodrick for sample preparation for the O-link assay with support from the NIH (K23MH119566), Krzysztof Hyrc for imaging and scanning services at the Hope Center Alafi Neuroimaging Laboratory (supported by NIH grants S10RR027552 and S10OD032121), Kymberli May and the WashU DDRCC Tissue Analysis & Imaging Core for tissue embedding and sectioning (supported by NIH grant P30DK052574), and the Center for Cellular Imaging (with support from NIH grant OD021629).

## Funding

National Institutes of Health grant DK131107 (JIG)

Gates Foundation INV-033564 (JIG)

Washington University School of Medicine Personalized Medicine Initiative (JIG)

National Institutes of Health grant K99HD122916 (KMP)

National Institutes of Health grant T32DK007130 (ZC)

National Institutes of Health grant F30HD115307 (RC)

American Gastroenterology Association Research Foundation’s AGA-Ironwood Fellowship-to-Faculty Transition Award AGA2024-32-02 (ZC)

Helen Hay Whitney Foundation fellowship (KMP)

## Author contributions

Conceptualization: KMP, ZLC, TA, MJB, JIG

Methodology: KMP, ZLC, MM, TA, MJB, JIG

Investigation: KMP, ZLC, MSH, MMR, MM, RC, RS, AHMR, SAS, SD, SMF, MAG, KAH, AMR, HL, RK, AEB, CK, BBB, AR, BB, DK, JF, MC

Formal analysis: KMP, ZLC

Visualization: KMP, ZLC

Funding acquisition: JIG

Project administration: KMP, ZLC, MC, TA, MJB, JIG

Supervision: JIG

Writing – original draft: KMP, ZLC, RC, MJB, JIG

Writing – review & editing: KMP, ZLC, JIG

## Data and materials availability

DNA sequencing, bulk RNA-seq, and snRNA-seq data have been deposited in NCBI’s Sequence Read Archive (SRA) under project number PRJNA1489488 and will be made publicly available prior to publication. This study did not use any original code. Additional information required to reanalyze the data reported in this paper is available from the lead contact upon request.

---

**Extended Data Fig. 1.**
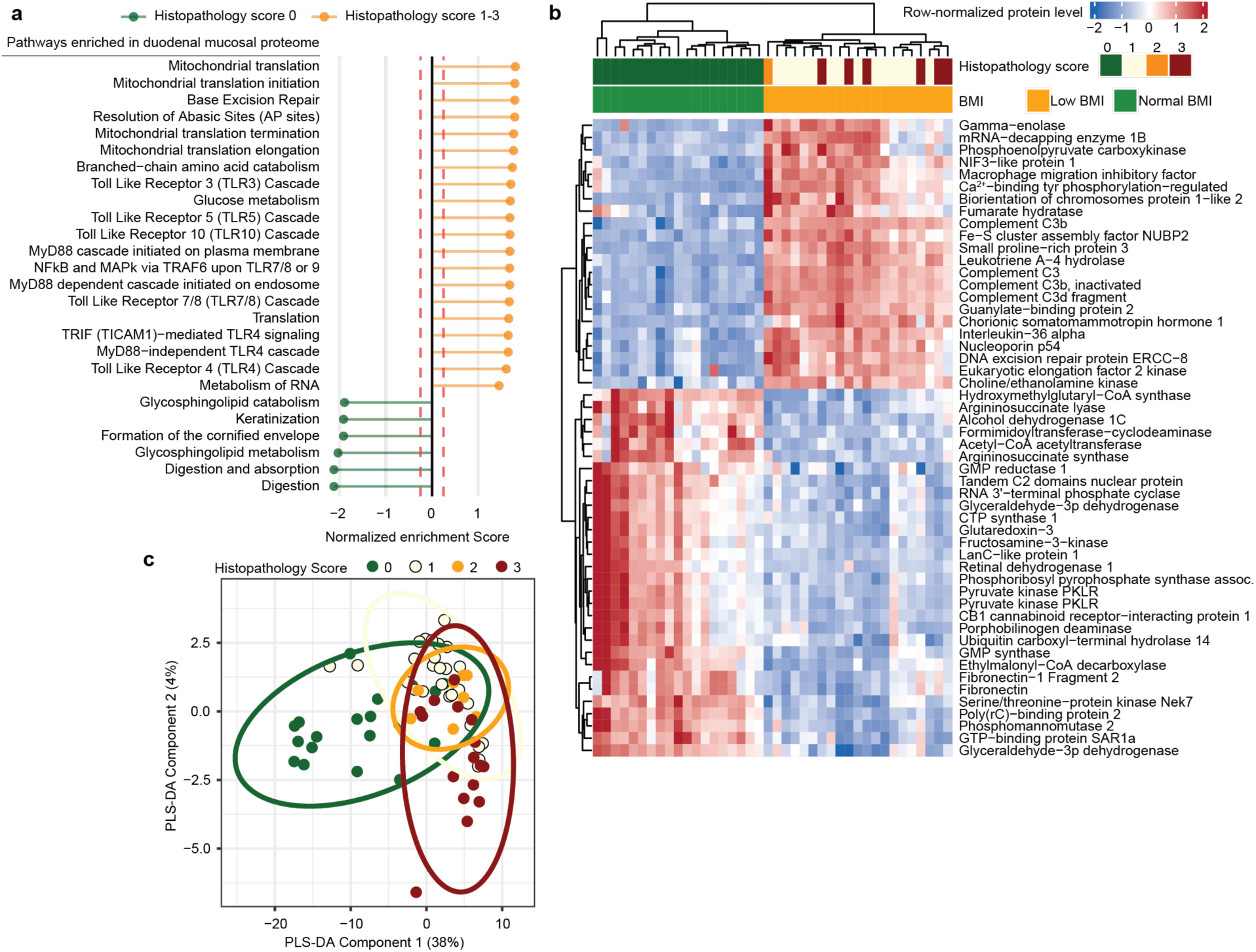
Duodenal mucosal and plasma proteomic analysis of undernourished women with EED compared to healthy controls. **a,** Reactome pathways enriched (GSEA) in the duodenal mucosal proteome of women with EED (histopathology score 1-3, n=59 women) compared to healthy women (histopathology score 0, n=19 women). **b,** Plasma proteins whose levels were significantly different between undernourished women with EED (n=22) and healthy controls (n=19). Values are row-normalized log_10_ values; (*limma P-*adj<0.01 and log_2_ fold difference>0.5 for inclusion in the heatmap). **c,** Dimensionality reduction of bacterial composition within duodenal aspirates, colored by histopathology score (Partial least squares discriminant analysis, PLS-DA). Ellipses denote 95% confidence intervals (n=80 women total; n=20 histopathology score 0, n=36 score 1, n=8 score 2, n=16 score 3).

**Extended Data Fig. 2.**
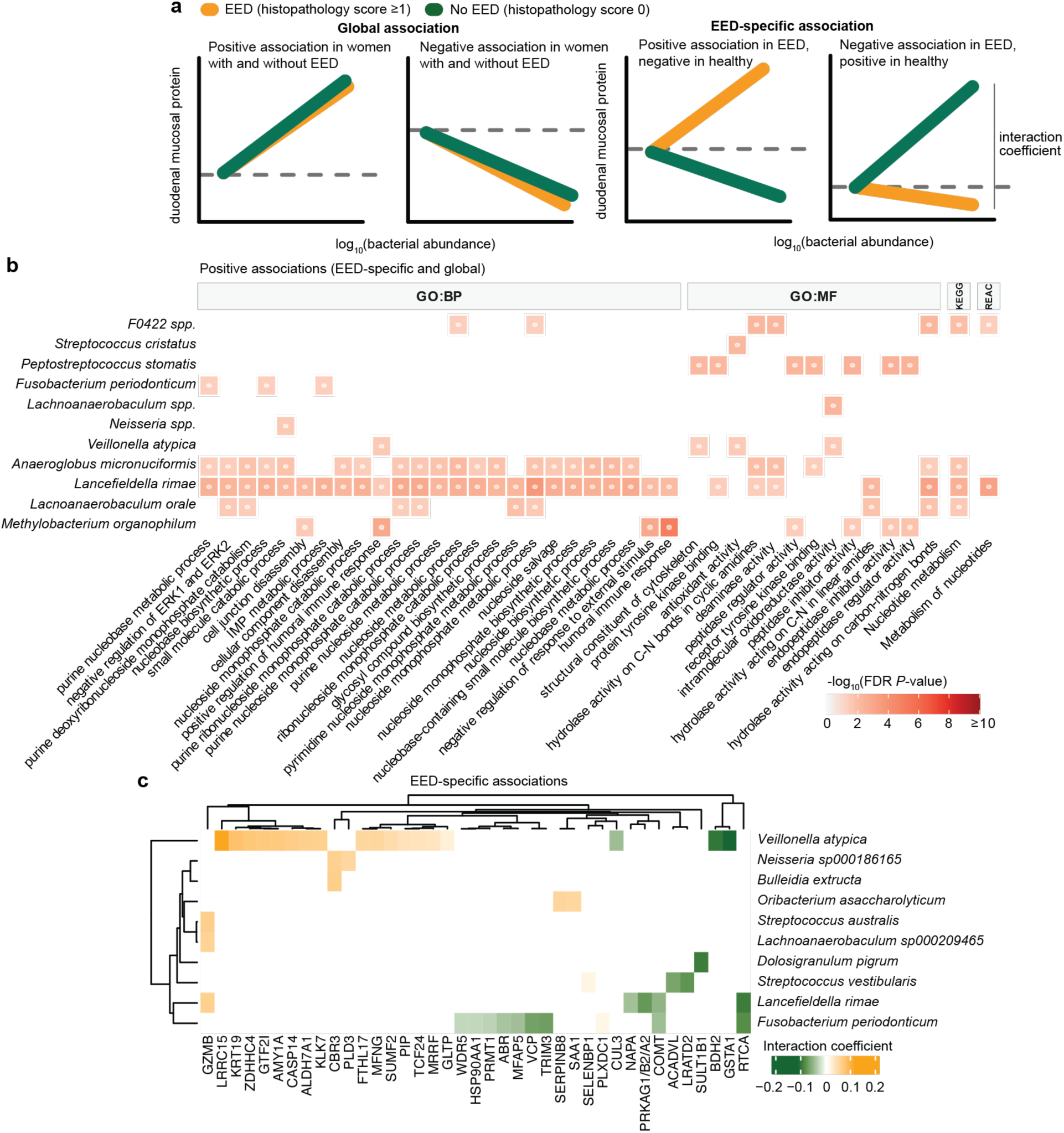
Associations between bacterial taxa in duodenal aspirates and duodenal mucosal proteins. **a,** Schematic representation of linear relationships between bacteria and proteins. **b,** Over-representation analysis performed on duodenal mucosal proteins whose levels were significantly positively linearly related to the absolute abundances of bacteria in aspirates (40 most significant categories shared amongst two taxa are shown). Abbreviations: ‘GO’, Gene ontology; ‘BP’, Biological Process’; ‘MF’, Molecular Function’; ‘KEGG’, Kyoto Encyclopedia of Genes and Genomes; ‘REAC’, Reactome. **c,** Bacteria-protein relationships whose slopes were significantly different between women with EED compared to those without (EED-specific associations). A positive interaction coefficient indicates a significantly more positive association in women with EED; a negative interaction coefficient, more negative in women with EED.

**Extended Data Fig. 3.**
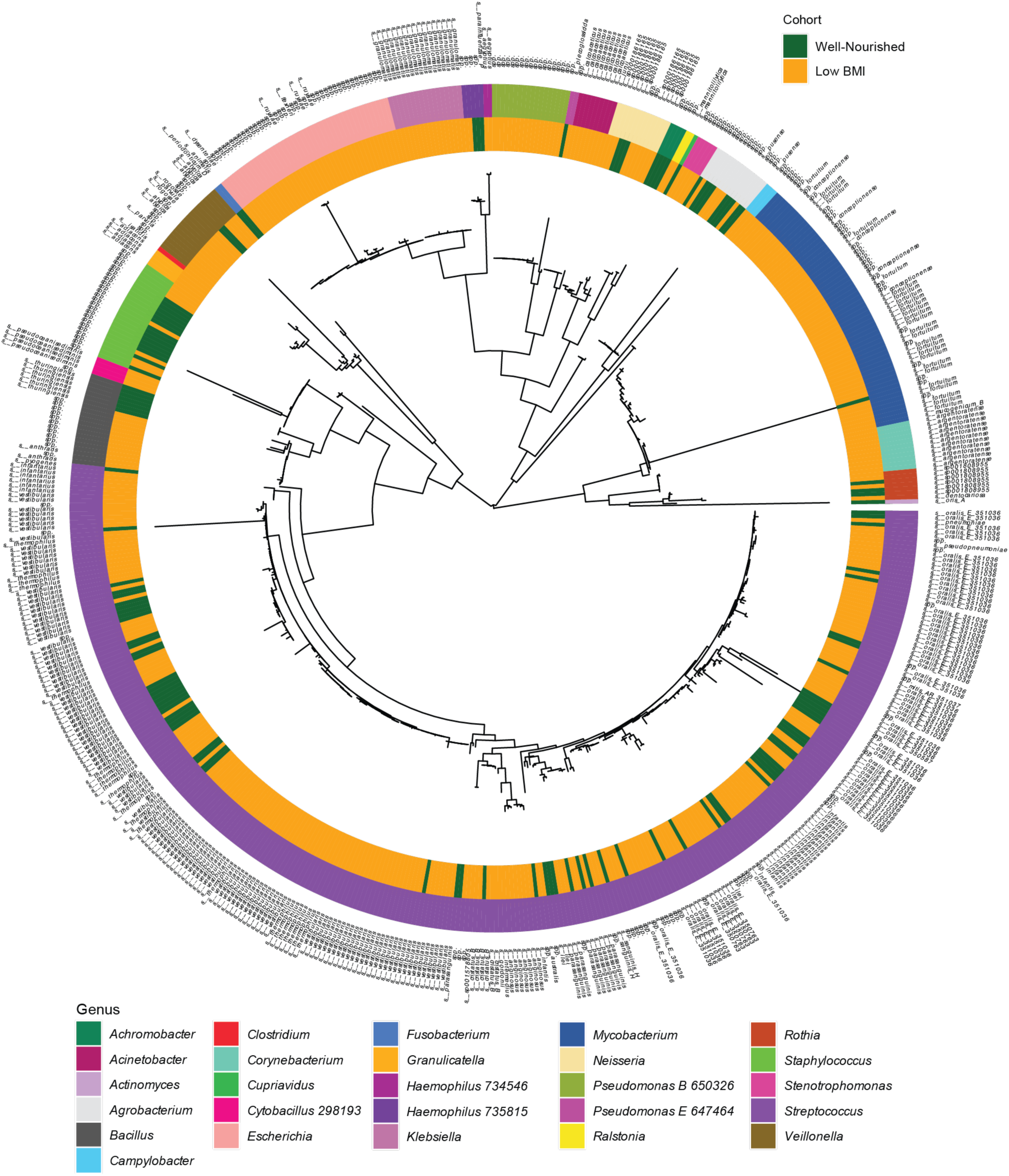
Bacteria cultured from duodenal aspirates obtained from Bangladeshi women. Phylogenetic tree of the 480 isolates cultured from members of the Low BMI cohort (orange) and 125 isolates from women in the Well-nourished cohort (green). The outer colored ring indicates genus-level assignments; the text denotes species-level assignments.

**Extended Data Fig. 4.**
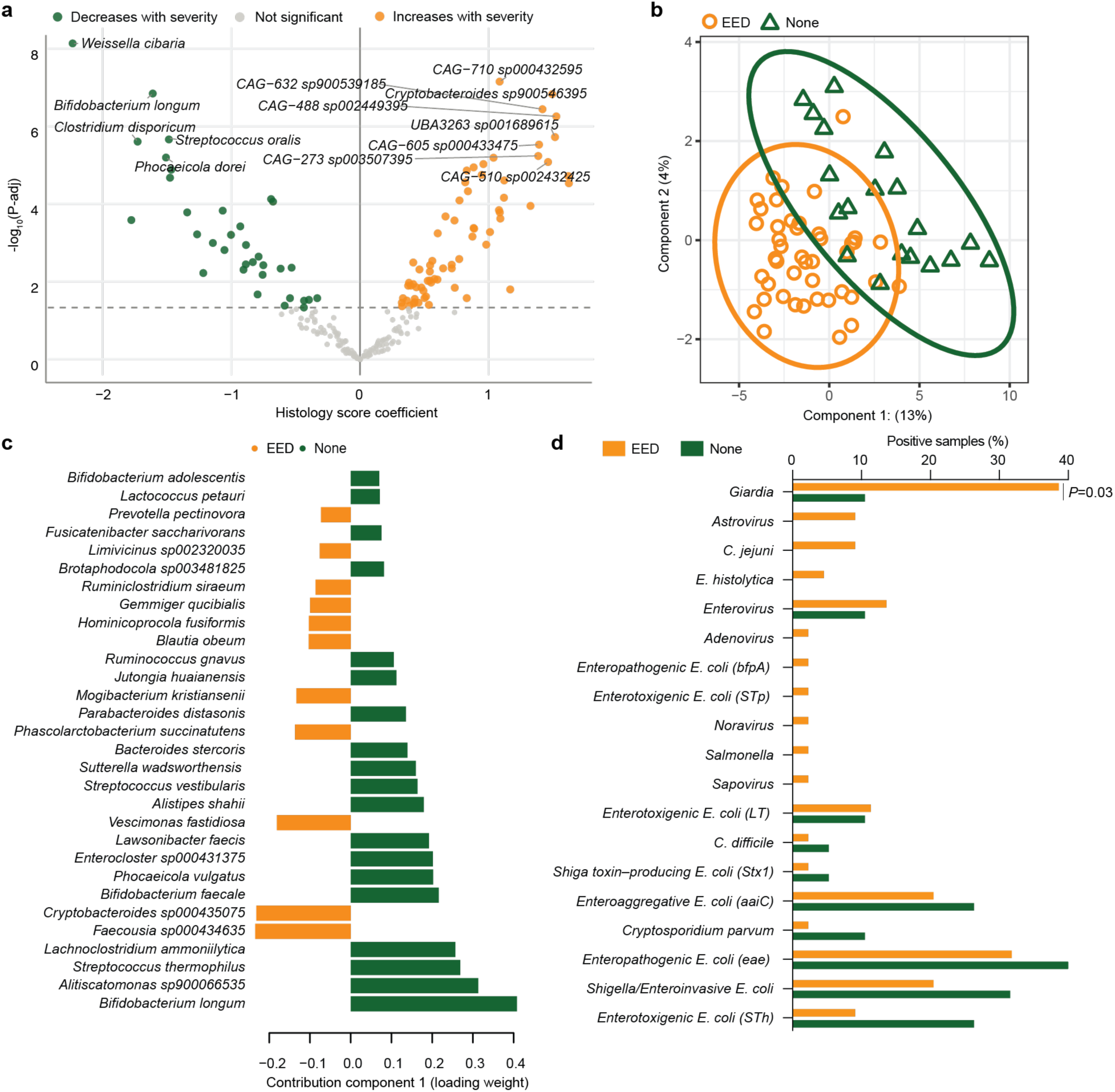
Fecal microbiota in women with EED. **a,** Bacteria whose relative abundances are significantly associated with histopathology score higher (orange, increase with histopathology score; green, decrease with histopathology score). **b,** Sparse partial least squares discriminant analysis (sPLS-DA) of the relative abundance of bacterial species in fecal samples from women with histopathologic evidence of EED compared to healthy controls. Ellipses denote 95% confidence intervals. **c**, Fecal bacteria whose relative abundance predicts disease status (sPLS-DA analysis, as in **b**). **d,** Prevalence of common enteropathogens in feces of undernourished and healthy women detected with a microfluidics qPCR-based platform (Fischer’s exact).

**Extended Data Fig. 5.**
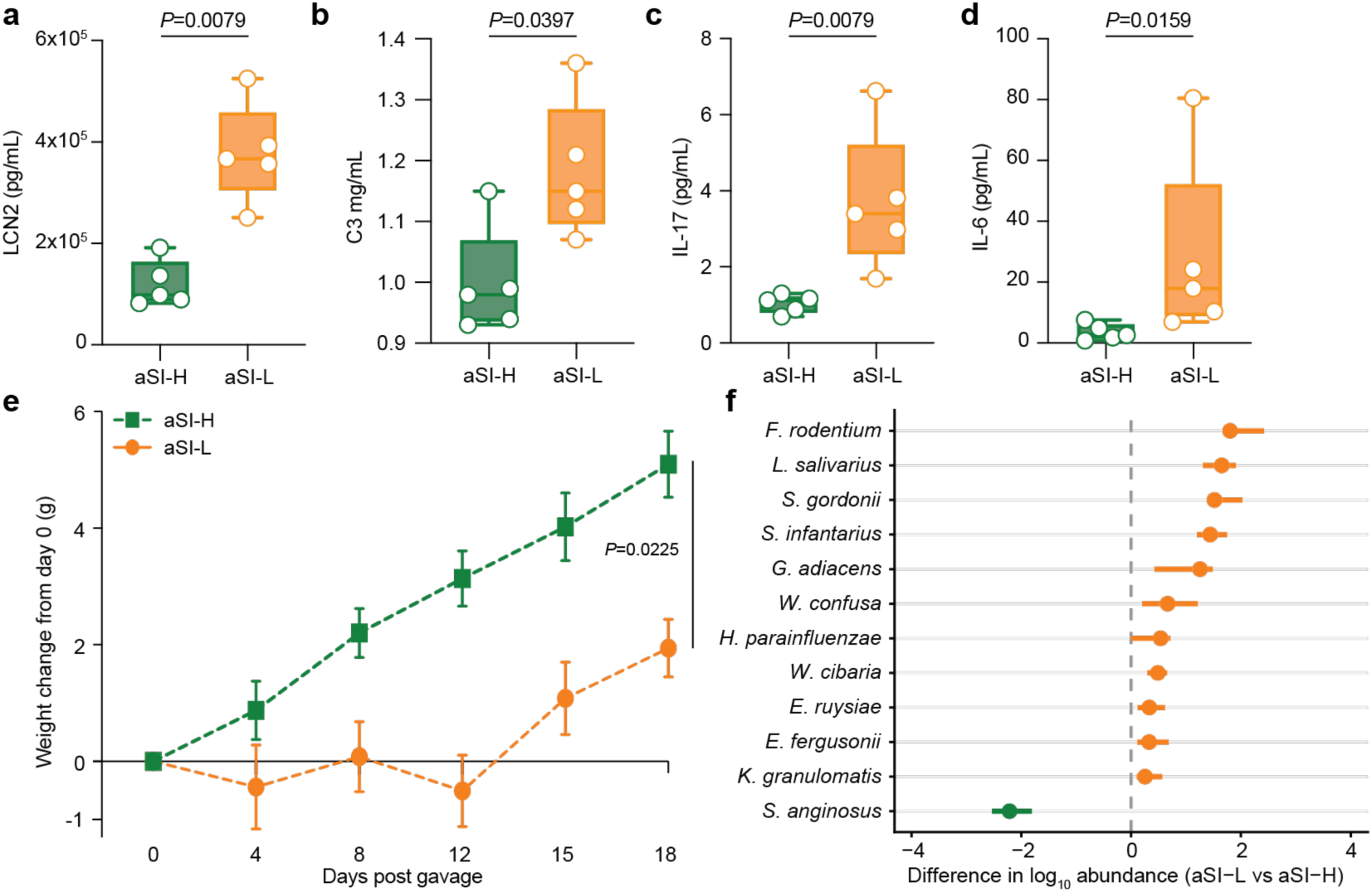
Evaluating consortia of bacteria derived from undernourished or healthy women in gnotobiotic mice. **a-d,** Serum levels of LCN2 (**a**), complement protein C3 (**b**), IL-17A (**c**) and IL-6 (**d**) after an 18-day colonization in mice harboring the aSI-L or aSI-H consortia (Wilcoxon rank-sum tests; boxes denote mean and inter-quartile range). **e,** Post-colonization weight gain (linear regression F_(1,48)_=5.556; mean values ± s.d. are shown). **f,** Statistically significant differences in absolute abundances of bacteria (Benjamini-Hochberg corrected Wilcoxon rank-sum tests; mean ± s.d.). For all panels, n=5 mice/group.

**Extended Data Fig. 6.**
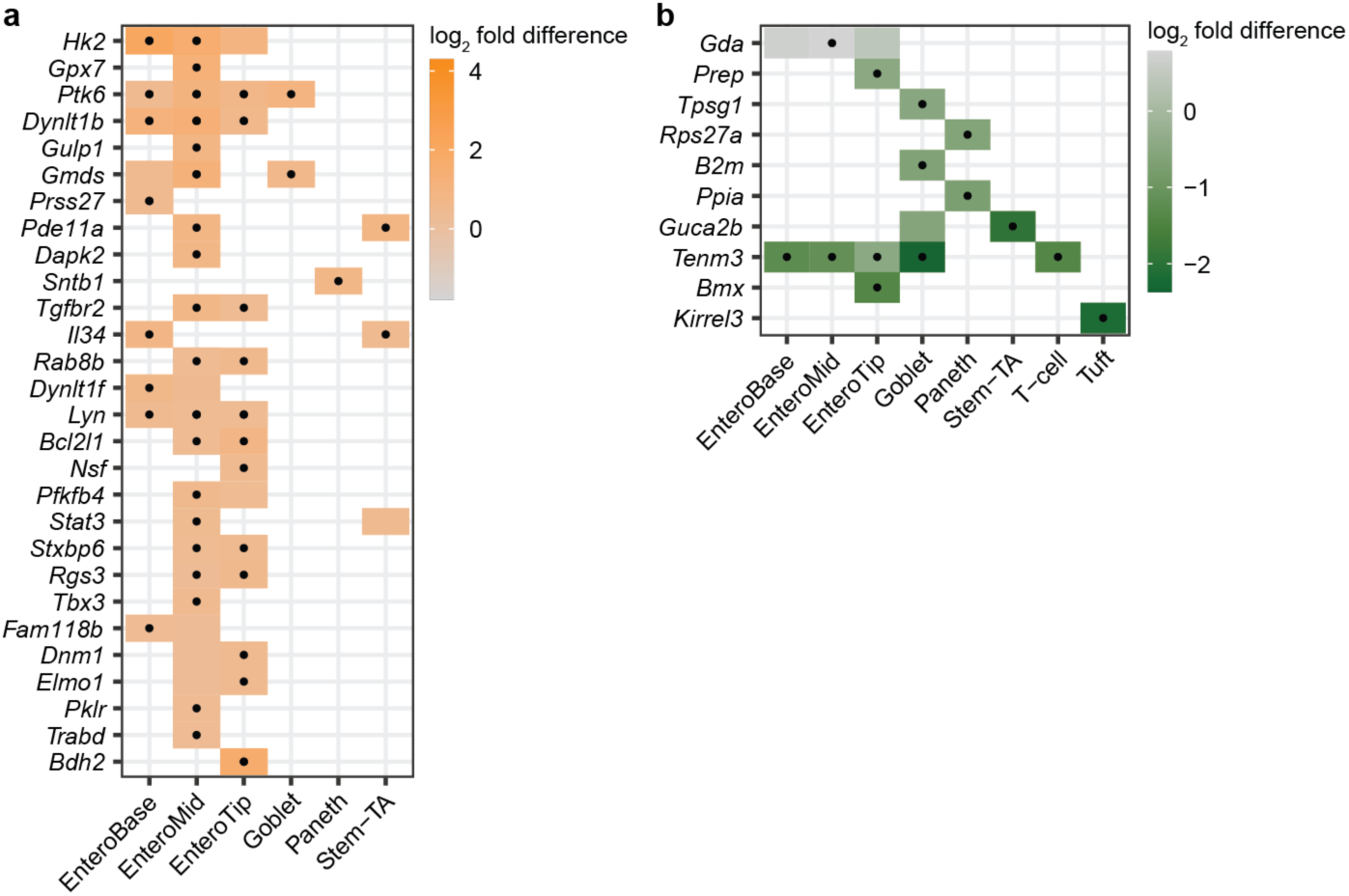
Differential expression of mouse genes that are homologous to human duodenal mucosal proteins with significantly different levels between women with and without EED. Results from snRNA-Seq analysis of duodenal segments harvested from aSI-L and aSI-H adult female animals. **a,** Genes that were significantly more highly expressed (*DESeq2* Wald test, adjusted *P*-value < 0.05) in cell types within the duodenums of aSI-L compared to aSI-H adult female mice that are homologues of human proteins whose levels were significantly higher in women with EED (histopathology score ≥1) compared to healthy women without EED (histopathology score 0, *limma* adjusted *P*-value < 0.05). **b,** Genes that were significantly more highly expressed (*DESeq2* Wald test, adjusted *P*-value < 0.05) in aSI-H compared to aSI-L adult female mice that correspond to human proteins whose levels were significantly higher in healthy women (histopathology score 0) compared to women with EED (histopathology score 1-3, *limma* adjusted *P*-value < 0.05). For both panels, cell types from the single nucleus RNA-Seq dataset were included if more than 100 nuclei were present, n=4 mice/group, • *P*-adj < 0.05. Abbreviations: Stem-TA, intestinal stem cell and transit amplifying cells; EnteroBase, EnteroMid, EnteroTip, enterocytes from villus base, mid-villus, and villus tip.

**Extended Data Fig. 7.**
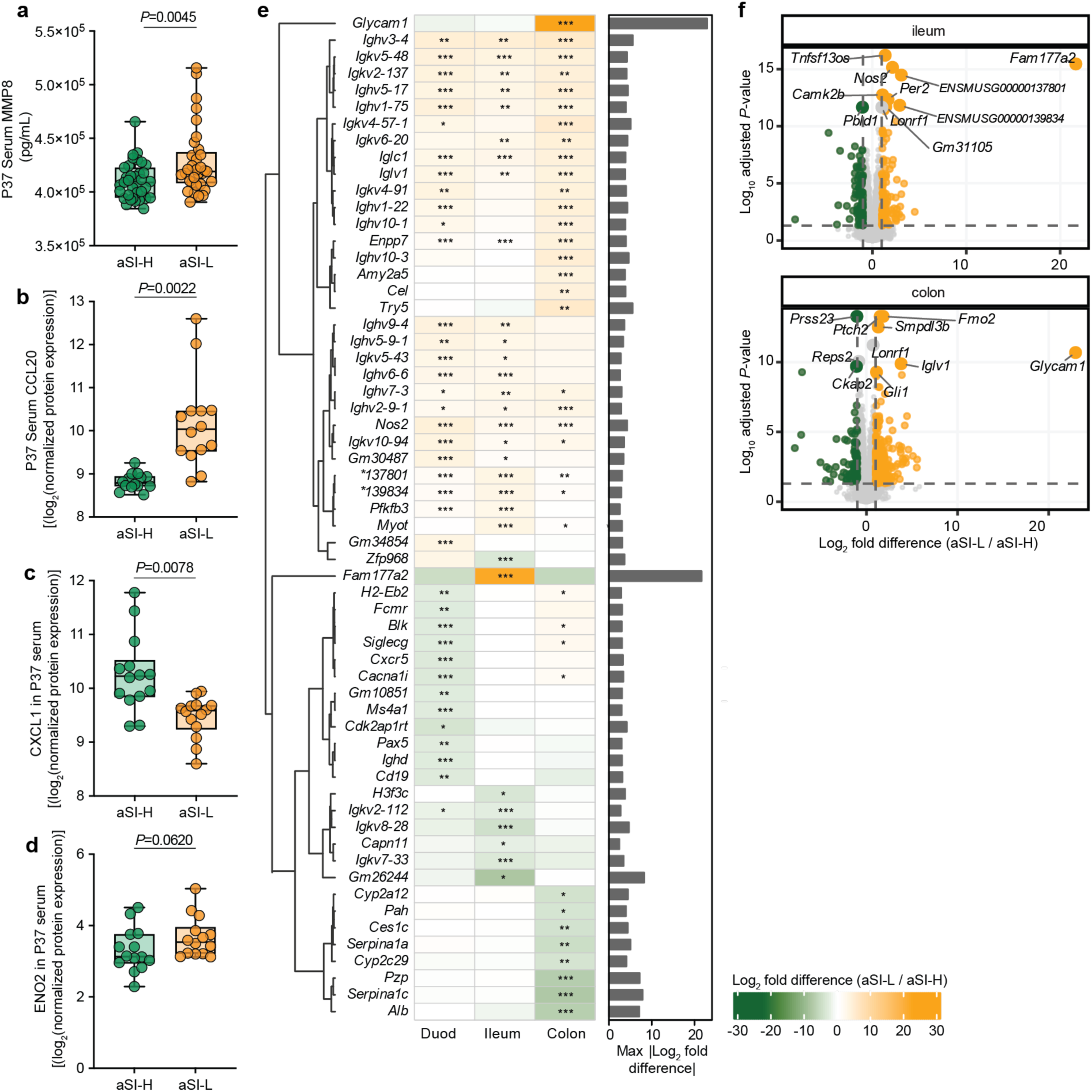
Systemic and intestinal inflammation in P37 offspring. **a-d,** Serum levels of MMP8 (**a**), CCL20 (**b**), CXCL1 (**c**) and ENO2 (**d**). CCL20, CXCL1 and ENO2 were quantified by O-link and reported as within-assay normalized protein expression. **e,f,** Genes most significantly differentially expressed between aSI-L (orange, positive log_2_-fold difference) and aSI-H (green, negative) offspring in the duodenum, ileum, and colon. **e,** Each column of the heatmap shows pairwise comparisons for each intestinal segment (*DESeq2* Wald test, * *P*-adj < 0.05, ** *P*-adj < 0.01, *** *P*-adj < 0.001). Bar chart annotation shows the maximum absolute log_2_-fold difference for each gene. **f,** Volcano plots depicting the most significantly expressed genes in the ileum and colon (*DESeq2* Wald test). For **a**, n=35 mice/group, unpaired two-sided t-test. For **b**-**d**, Benjamini Hochberg-corrected Wilcoxon rank-sum test, n=14 animals/group.

**Extended Data Fig. 8.**
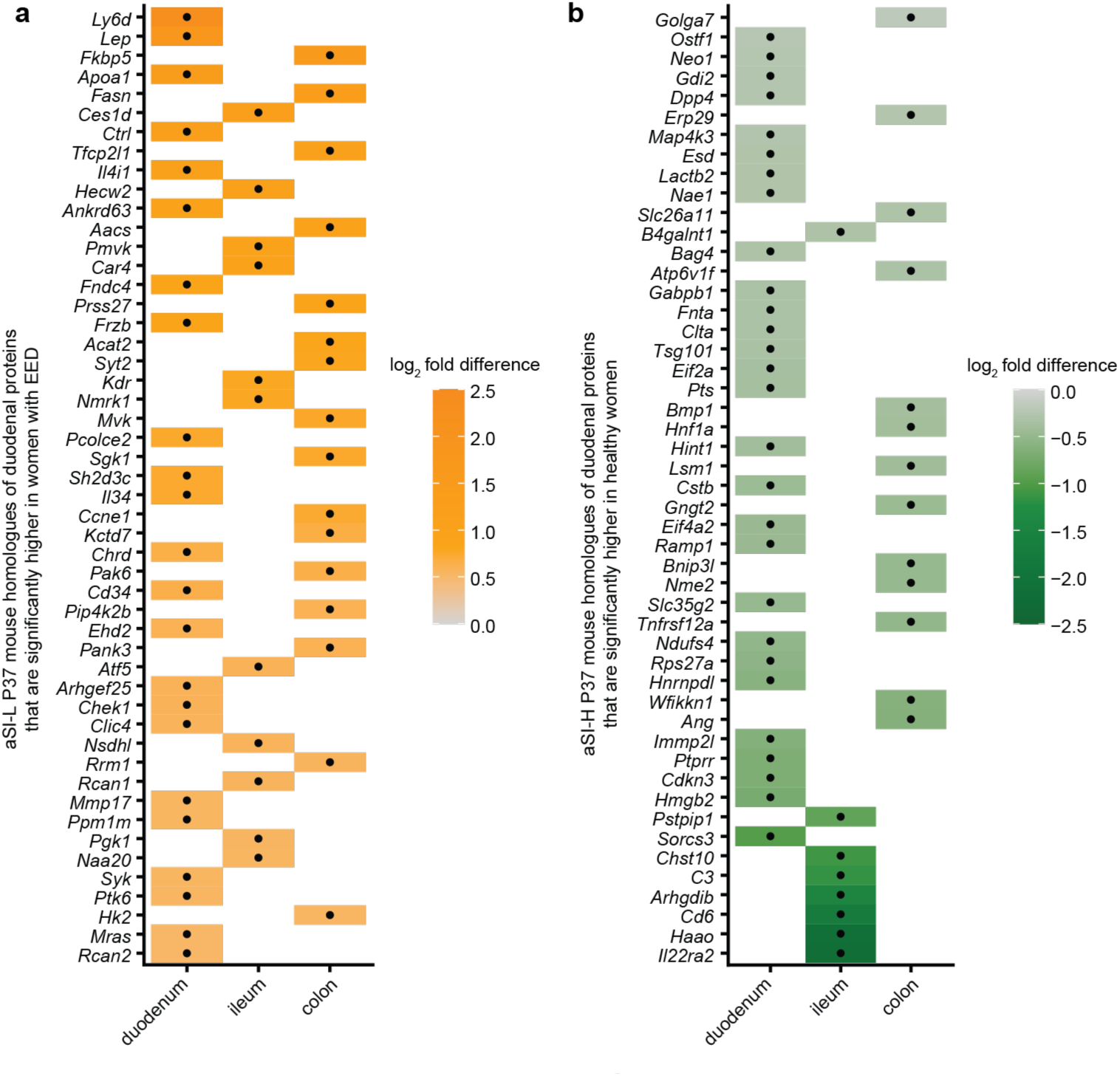
Expression in P37 intestinal tissue of mouse genes homologous to proteins whose levels were significantly different between women with and without EED. **a,b** Mouse genes that were significantly more highly expressed in aSI-L compared to aSI-H P37 offspring (**a**) or aSI-H compared to aSI-L (**b**) P37 offspring (*DESeq2* Wald test, • *P*-adj < 0.05).

**Extended Data Fig. 9.**
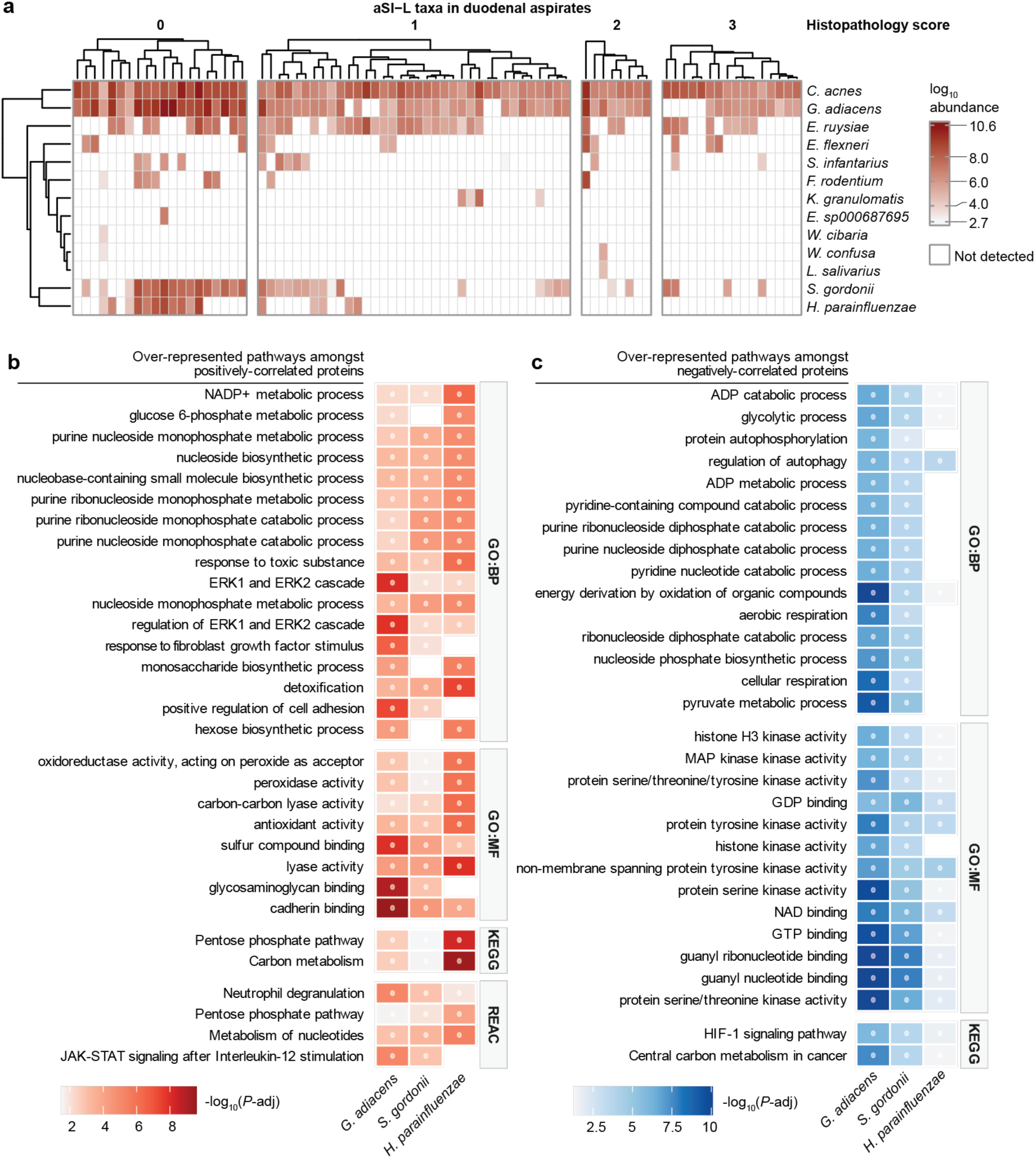
Correlations between aSI-L taxa in duodenal aspirates with duodenal mucosal proteins. **a,** Log_10_-transformed absolute abundances of aSI-L taxa in duodenal aspirates (n=80 women; aspirates binned according to histopathology score). **b,c** Enrichment analysis was performed on the sets of duodenal proteins that were significantly positively (**b**) or negatively (**c**) correlated with the absolute abundances of *G. adiacens, S. gordonii* and *H. parainfluenzae* in the aspirates of women (n=78 women; 30 most significant categories are included; color denotes -log_10_-transformed FDR-corrected P-values, •, *P*-adj < 0.05). Abbreviations: ‘GO’, Gene ontology; ‘BP’, Biological Process’; ‘MF’, Molecular Function’; ‘KEGG’, Kyoto Encyclopedia of Genes and Genomes; ‘REAC’, Reactome.

**Extended Data Fig. 10.**
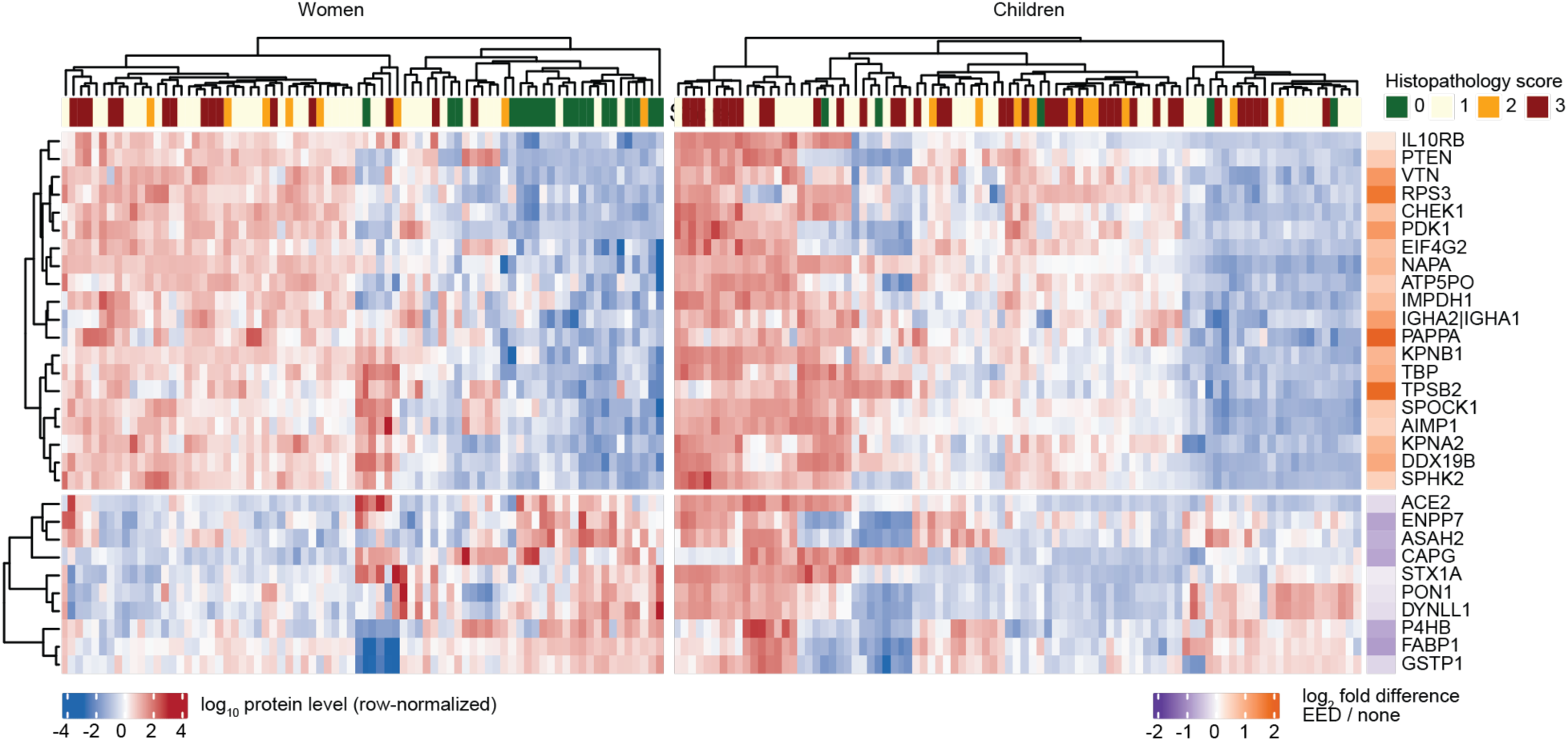
Duodenal mucosal proteins whose levels differ between women and children with and without EED. Duodenal mucosal proteins that were the most significantly different between women and children with severe EED (histopathology score 3, n=15/78 women in the current study, 37/89 children in the BEED study who were undernourished and underwent EGD^14^) compared to those without evidence of enteropathy (histopathology score 0; n=19/78 women, 5 of the 89 children^14^; *limma* empirical Bayes moderation). Log_10_-transformed protein levels are row-normalized within each cohort; the top annotation denotes histopathology score, and the right row annotation shows the log_2_ fold-difference across both women and children (n=78 biopsies from women, n=89 from children).

## SUPPLEMENTARY TABLES

**Supplementary Table 1. Biospecimens acquired and proteomics analysis of clinical cohorts. a,** Metadata and biospecimens collected from human studies. **b,** (i) Duodenal mucosal proteins that differed significantly between women with EED (histopathology score ≥ 1) and without EED (histopathology score 0) (*limma* with empirical Bayes moderation). (ii) Reactome pathways that were significantly enriched in the duodenal mucosal proteome of women with EED compared to those without histologic evidence of EED (from i, gene-set enrichment analysis performed with *clusterProfiler*). **c,** Plasma proteins that differed significantly between undernourished women with EED (histopathology score ≥ 1) and without EED (histopathology score 0) (*limma* with empirical Bayes moderation). **d,** Proteins that were significantly elevated in both duodenal biopsies and plasma of women with histopathologic evidence of EED (*P*-adj<0.1 in both comparisons).

**Supplementary Table 2. Duodenal microbiota of women with EED. a,** Absolute abundances of bacterial taxa in duodenal aspirates (log_10_-transformed 16S rRNA counts per gram aspirate). (i) Present in at least n=10/80 duodenal aspirate samples. (ii) All bacteria detected. **b,** Results of zero-inflated Gaussian mixture model testing for significant associations between the absolute abundance of bacterial taxa in duodenal aspirates and histopathology score. **c,** Duodenal bacterial taxa whose absolute abundance differed between (i) histopathology scores and (ii) women with (histopathology score ≥ 1) or without (histopathology score 0) evidence of EED (linear models on log_10_-transformed absolute abundances).

**Supplementary Table 3. Analyses associating duodenal bacterial taxa and duodenal mucosal proteins in women. a-b,** Duodenal mucosal proteins (**a**) and bacteria in duodenal aspirates (**b**) that were predictive of EED status (histopathology score ≥ 1 vs. 0 (Data Integration Analysis for Biomarker discovery using Latent variable approaches for Omics studies, ‘DIABLO’). **c,** Results of linear models relating absolute abundances of taxa in duodenal aspirates to duodenal mucosal protein levels. **d-e,** Functional enrichment analysis performed on duodenal mucosal proteins that were positively (**d**) or negatively (**e**) related to absolute abundance of bacterial taxa (*gProfiler2* over-representation test).

**Supplementary Table 4. Fecal microbiota of women with EED. a,** Relative abundance (percent) of bacterial taxa in fecal samples. (i) Present in at least n=9/63 fecal samples. (ii) All bacteria detected. **b,** Fecal bacteria whose relative abundances (i) were significantly associated with histopathology score and (ii) differed significantly by disease status (histopathology score ≥ 1 compared to 0) (zero-inflated Gaussian mixture model). **c,** Enteric pathogens detected in DNA extracted from fecal samples using a microfluidics-based qPCR platform.

**Supplementary Table 5. Bacteria cultured from duodenal aspirates of women in the Low BMI and Well-nourished cohorts. a,** Summary of the number of isolates obtained per species. **b,** Isolates with their 16S rRNA sequences (Sanger) and taxonomic identification (GreenGenes; 2024 release).

**Supplementary Table 6. Quantification of protein biomarkers and bacterial abundance in first tests of aSI-L and aSI-H consortia. a,** Quantification of serum biomarkers in (i) just-weaned mice colonized with pooled culture collections from Low BMI and Well-nourished women and (ii) mice colonized with cecal contents obtained from mice harboring intact culture collections from Well-nourished (NBMI) or Low BMI (LBMI) donors that produced high (hi) or low (lo) levels of inflammation based on rank sum of the biomarkers. **b,** Changes in body mass over time in mice described in **a**(ii). **c,** Quantitation of bacterial taxa for mice described in **a** (log_10_-transformed 16S rRNA counts per g cecal contents). **d,** Differences in the absolute abundances of bacteria in aSI-L compared to aSI-H mice (Benjamini Hochberg-adjusted Wilcoxon rank-sum tests).

**Supplementary Table 7. Small intestinal histomorphometric and snRNA-Seq plus proteomic analyses of adult female aSI-L and aSI-H mice. a,** Histomorphometric analysis (10 measurements/small intestinal segment from n=4-5 mice/treatment group). Statistical significance assessed by linear mixed-effects models (*Measure ∼ Microbiota + (1 | Mouse_ID*)). **b**-**d,** ‘Pseudo-bulk’ differential expression analysis of snRNA-Seq data acquired from the duodenum (**b**), jejunum (**c**), and ileum (**d**) of adult female aSI-L mice (positive log_2_-fold change) and aSI-H mice (negative log_2_-fold change) (Benjamini Hochberg-adjusted Wald test using *DESeq2*). **e,** Quantification of protein biomarkers in serum (i, ELISA) and intestinal tissue (i, Luminex and ii, immunofluorescence) of aSI-L and aSI-H adult female mice. **f,** Immune cells quantified by flow cytometry in the small intestinal and colonic lamina propria of aSI-L and aSI-H adult females. **g,** Antibodies used for flow cytometry in f. **h,** Quantification of FITC-conjugated 4 kDa dextran in the serum of aSI-L and aSI-H adult females.

**Supplementary Table 8. Serum proteomic, bone micro-computed tomography and intestinal bulk RNA-Seq analyses of P37 offspring of aSI-L and aSI-H dams. a,** Serum proteins quantified with ELISAs (i) and the Olink Target 96 Mouse Exploratory platform (ii, Benjamini Hochberg-adjusted Wilcoxon rank-sum tests). **b,** Micro-computed tomography measurements of femurs: (i), cortical bone tissue; (ii), trabecular bone tissue (two-sample two-tailed t-tests). **c,** Body weights. **d,** Differential gene expression between aSI-L offspring (positive log_2_-fold change) and aSI-H offspring (negative log_2_-fold change) (Benjamini Hochberg-adjusted Wald tests using *DESeq2*, conducted separately for each tissue). **e,** Results of GSEA performed on differentially expressed genes in intestinal tissue (as in D). **f,** Differential expression of genes encoding mouse homologs of duodenal mucosal proteins that were significantly higher in women with EED (as in **Supplementary Table 1b**). (i), Mouse homologs of proteins that were higher in women with EED and more highly expressed in the intestinal tissue of aSI-L offspring. (ii), Mouse homologs to proteins that were diminished in women with EED compared to healthy women and more highly expressed in the intestinal tissue of aSI-H offspring. **g,** Bacterial abundance in adult females and P37 offspring (log_10_-transformed 16S rRNA counts per g cecal contents).

**Supplementary Table 9. Correlations between bacterial taxa associated with inflammation in pre-clinical mouse models and duodenal mucosal proteins in women. a,** Correlations between the absolute abundances of aSI-L bacteria in duodenal aspirates of women and levels of proteins quantified in their duodenal mucosal biopsies (Spearman correlations were performed for bacteria that were present in greater than 10 women). **b,** Functional enrichment analysis (*gProfiler2* over-representation test) performed on duodenal mucosal proteins significantly correlated with aSI-L bacteria in aspirates (results from **a**). **c,** Duodenal mucosal proteins whose levels were significantly different between women and children with severe EED (histopathology score 3) compared to women and children without pathology (score 0). **d,** 16S rRNA sequence and taxonomic matches between bacteria cultured from children with EED that did (P37 cSI-I offspring) or did not (P37 cSI-N offspring) induce immunoinflammatory responses^16^ and bacterial taxa in duodenal aspirates of adult women in the current study. **e,** Correlations between the absolute abundances of bacteria cultured from children with EED detected in aspirates of women and levels of their duodenal mucosal proteins (FDR-adjusted Spearman correlations). (i), Bacterial taxa that induced immuno-inflammatory responses in P37 offspring (‘cSI-I’; ref^16^); (ii), Bacterial taxa that did not induce inflammation (‘cSI-N’; ref.^16^). **f,g,** GSEA (i, *clusterProfiler*) and over-representation tests (ii, *gProfiler2*) performed on duodenal mucosal proteins that were significantly correlated with the abundances of cSI-I (**f**) or cSI-N (**g**) bacterial taxa.

