## Supplementary Results for "Duodenal microbiota and associated environmental enteric dysfunction in undernourished Bangladeshi women and gnotobiotic mice"

### Duodenal bacterial taxa and duodenal proteins predictive of EED status

We utilized a supervised learning approach, 'Data Integration Analysis for Biomarker discovery using Latent variable approaches for 'Omics' (DIABLO; a 'block' sparse partial least-squares discriminant analysis), to find predictive bacterial and proteomic features of EED (**Supplementary Fig. 1a,b**)[53]. The overall balanced error rate for the DIABLO model was 0.0987. Duodenal mucosal proteins that were predictive of EED (histopathology score ≥1) included Tyrosine Kinase (SYK), which mediates inflammatory responses at the intestinal epithelium and multiple leukocyte lineages; Receptor for activated C kinase 1 (RACK1), which regulates E-cadherin and catenin complexes and regulates cell differentiation in the crypts, Ring Finger Protein 31 (RNF31), which activates NF-kB; as well as proteins involved in proliferation [Eukaryotic Elongation Factor 2 Kinase (EEF2K), Deoxyhypusine Hydroxylase (DOHH)] and DNA repair [8-Oxoguanine glycosylase (OGG1), Polynucleotide Kinase 3'-Phosphatase (PNKP)] (**Supplementary Fig. 1c, Supplementary table 3a)**. Osteoclast Stimulating Factor 1 (OSTF1), which is involved in cytoskeletal organization and cell signaling, was predictive of the absence of EED (**Supplementary Fig. 1c, Supplementary table 3a**). Duodenal bacteria most predictive of the absence of pathology included *Bulleidia timonensis*, *Lancefieldella rimae*, *Lachnoanaerobaculum orale*, *Anaeroglobus micronuciformis* and *Rothia aeria* (**Supplementary Figs. 1d** and **2, Supplementary table 3b**), all of which are considered commensal members of the oral microbiota. This approach integrates abundances of bacterial taxa in duodenal aspirates with levels of proteins in the duodenal mucosa to predict disease status, identifying features most strongly associated with the presence or absence of EED.

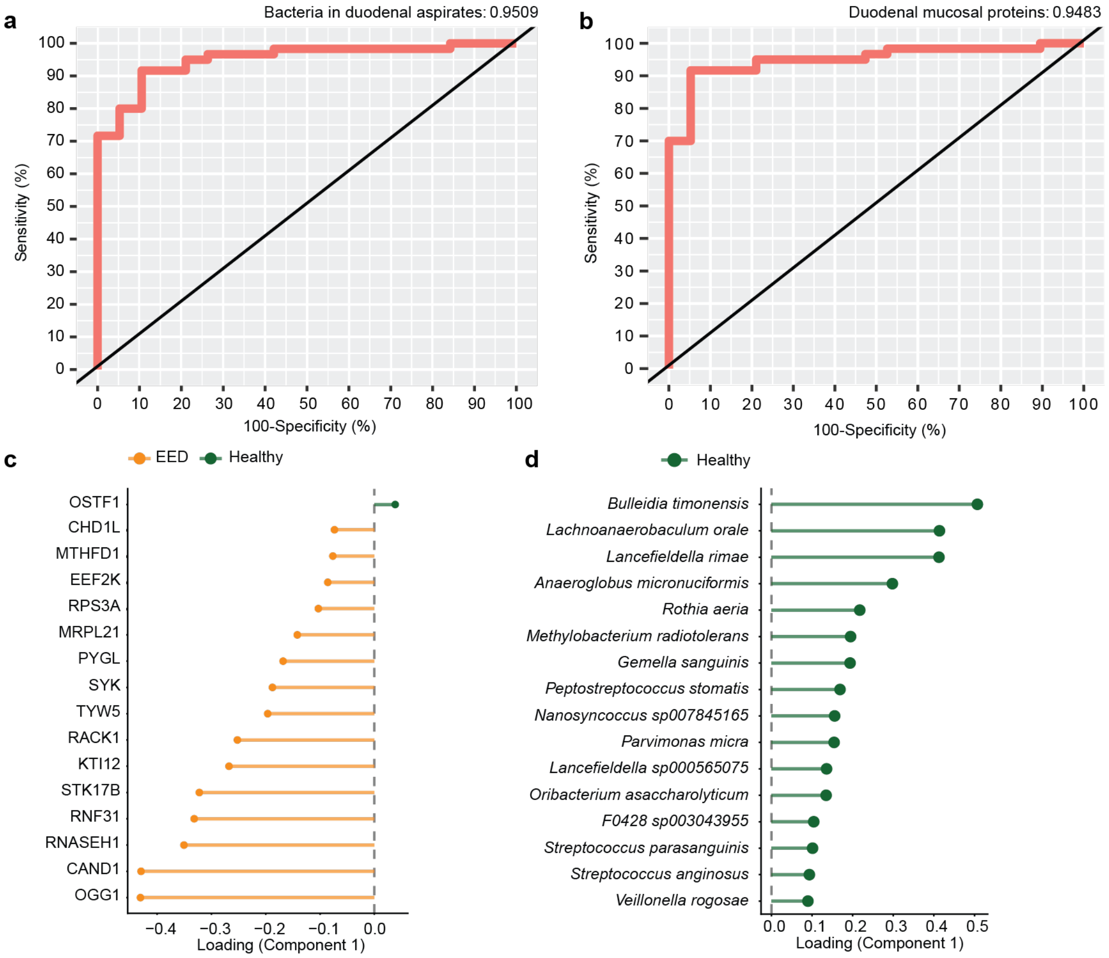


**Supplementary Fig. 1. Feature prediction for DIABLO analysis. a,b** Area Under the Receiver Operating Characteristic (AUROC) plots depicting the individual performance of the binary classification models for bacteria in duodenal aspirates (**a**) and proteins in duodenal biopsies (**b**), which were both part of the combined DIABLO model. **c,d** Proteins (**c**) and bacteria (**d**) in duodenal mucosal biopsies and aspirates, respectively, that predict disease status (EED: histopathology score 1-3, orange, n=59 out of 78 women; absence of detectable EED: green, histopathology score 0, n=19 out of 78 women. For **c** and **d**, the 16 most discriminatory features are included).

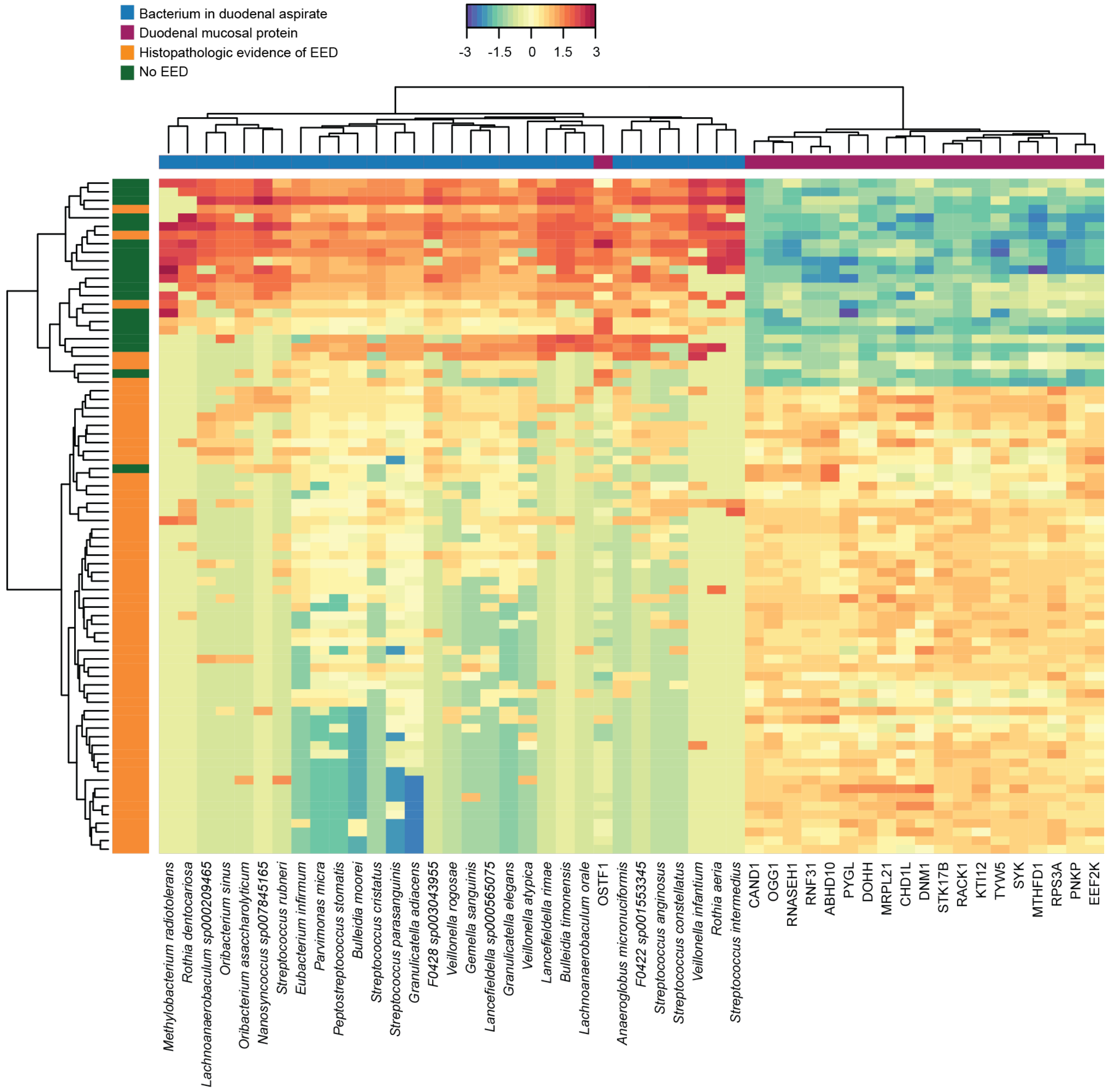


**Supplementary Fig. 2. Duodenal bacteria and mucosal proteins that predict disease status.** Heatmap depicting discriminatory features contributing to DIABLO analysis integrating absolute bacterial abundance in aspirates with the duodenal mucosal proteome of women with EED (histopathology score ≥1) compared to healthy controls (histopathology score 0).

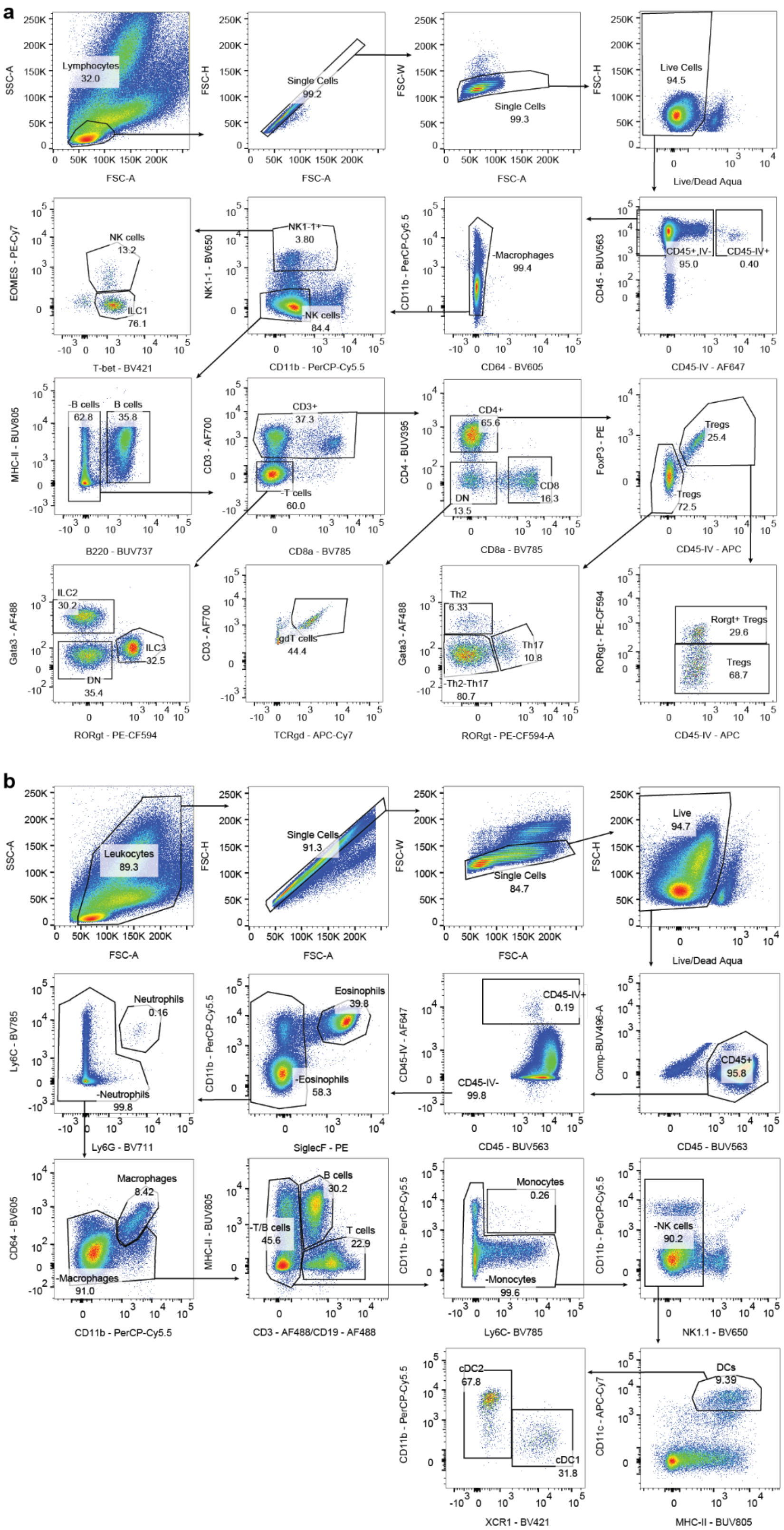


**Supplementary Fig. 3. Gating schemes used in flow cytometry analysis. a,** Gating strategy for identification of lymphoid populations. **b,** Gating strategy for identification of myeloid populations.
